# Genetic modification of inflammation and clonal hematopoiesis-associated cardiovascular risk

**DOI:** 10.1101/2022.12.08.22283237

**Authors:** Zhi Yu, Trevor P. Fidler, Yunfeng Ruan, Caitlyn Vlasschaert, Tetsushi Nakao, Md Mesbah Uddin, Taralynn Mack, Abhishek Niroula, J. Brett Heimlich, Seyedeh M. Zekavat, Christopher J. Gibson, Gabriel K. Griffin, Yuxuan Wang, Gina M. Peloso, Nancy Heard-Costa, Daniel Levy, Ramachandran S. Vasan, François Aguet, Kristin Ardlie, Kent D. Taylor, Stephen S. Rich, Jerome I. Rotter, Peter Libby, Siddhartha Jaiswal, Benjamin L. Ebert, Alexander G. Bick, Alan R. Tall, Pradeep Natarajan

## Abstract

Clonal hematopoiesis of indeterminate potential (CHIP) is associated with an increased risk of cardiovascular diseases (CVD), putatively via inflammasome activation. We pursued an inflammatory gene modifier scan for CHIP-associated CVD risk among 424,651 UK Biobank participants. CHIP was identified using whole exome sequencing data of blood DNA and modeled both as a composite and for common drivers (*DNMT3A*, *TET2*, *ASXL1,* and *JAK2*) separately. We developed predicted gene expression scores for 26 inflammasome-related genes and assessed how they modify CHIP-associated CVD risk. We identify *IL1RAP* as a potential key molecule for CHIP-associated CVD risk across genes and increased *AIM2* gene expression leading to heightened *JAK2*- and *ASXL1*-associated CVD risks. We show that CRISPR-induced *Asxl1* mutated murine macrophages have a particularly heightened inflammatory response to AIM2 agonism, associated with an increased DNA damage response, as well as increased IL-10 secretion, mirroring a CVD protective effect of *IL10* expression in *ASXL1* CHIP. Our study supports the role of inflammasomes in CHIP-associated CVD and provides new evidence to support gene-specific strategies to address CHIP-associated CVD risk.

## Introduction

Clonal hematopoiesis (CH) of indeterminate potential (CHIP) is the age-related acquisition and expansion of somatic mutations of genes frequently mutated in hematologic malignancies (e.g., *DNMT3A, TET2*, *ASXL1*, or *JAK2*) (1) detected from sequencing blood DNA among asymptomatic individuals. CHIP is common among older adults, affecting at least 1 in 10 adults over 70 years (2–5). CHIP is associated with an increased risk of hematologic malignancy and all-cause mortality (3, 4), as well as a range of cardiovascular diseases (CVD) (6–10). Recent evidence, primarily from murine and cell-based studies, suggests that dysregulated inflammation may be a key contributor to the augmented risk of CVD conferred by certain CHIP mutations (6, 11–14).

Heightened interleukin (IL)-1β signaling, a key inflammatory pathway, promotes the development of CHIP-associated atherosclerosis in *Tet2* CHIP as initially disclosed largely by murine studies (6, 11). Inhibition of the NOD-, LRR- and pyrin domain-containing protein 3 (Nlrp3) inflammasome abrogates accelerated atherosclerosis in atherogenic mice with hematopoietic *Tet2* deficiency versus wild-type (11, 15). In humans, CHIP is associated with increased gene expression and circulating concentrations of NLRP3 downstream products, particularly in the context of *TET2* CHIP (16–18). Humans harboring *IL6R* p.Asp358Ala, a common variant known to disrupt *IL6R* and associate with modestly reduced CVD risk reduction in the general population, had greater reductions in CVD risk among individuals with *DNMT3A* or *TET*2 CHIP mutations versus those without (7). However, recent murine work indicates that different CHIP genes may confer CVD risk differentially. For example, among atherogenic transgenic mice expressing *Jak2^VF^*, bone marrow genetic deficiency of the absent in melanoma 2 (*Aim2*) inflammasome mitigated atherosclerotic lesion development (15). Whether these findings extend to humans is currently not well understood. In general, the range of inflammatory cytokines differentially influencing CVD risk by CHIP genes in humans requires further study. Prioritization by human genetics may yield or bolster new approaches to CVD precision medicine (19).

To overcome risks of confounding from biomarker correlation analyses, we leveraged genetics to pursue a broader inflammatory gene modifier scan for CHIP -associated CVD among 424,651 UK Biobank participants with blood DNA exome sequencing for CHIP genotyping, array-derived genome-wide genotyping for transcriptomic imputation, and baseline and incident clinical outcomes. We developed predicted gene expression scores for genes related to the NLRP3 and AIM2 inflammasomes based on externally trained data and conducted independent validation. Then we assessed whether and to what extent the predicted gene expression modifies CHIP-associated CVD risk. Lastly, we validated a human genomics-based discovery in a murine model. Broadly, we demonstrate a systematic approach to prioritize potential therapeutic strategies for CHIP-associated disease.

## Results

### Baseline characteristics of the UK Biobank cohort

The schematic of this study is shown in **Figure 1**. Among the 417,570 unrelated participants enrolled in the UK Biobank study who underwent exome sequencing and were free of hematologic cancers and composite CVD events at baseline, the mean (SD) age was 56.3 (8.1) years, and 185,492 (44.4%) were men. We identified 25,784 (6.2%) individuals with CHIP mutations, with a mean (SD) age of 59.7 (7.1). Among participants with CHIP mutations, 92.6% have only one driver mutation; 14,297 (55.4%) had mutations in *DNMT3A*, 5,133 (19.9%) in *TET2*, and 2,436 (9.1%) in *ASXL1*. Two hundred and forty-eight participants (1.0%) had *JAK2* mutations, 222 (89.5%) of whom had *JAK2* p.V617F and 241 (97.2%) were large clones defined as a variant allele fraction (VAF) >10%. Consistent with previous reports, participants with CHIP versus those without were on average four years older, more likely to be White, have higher BMI, be ever smokers, and have a higher prevalence of cardiovascular comorbidities including hypertension, hyperlipidemia, and type 2 diabetes mellitus (**Table 1**).

**Figure 1.**
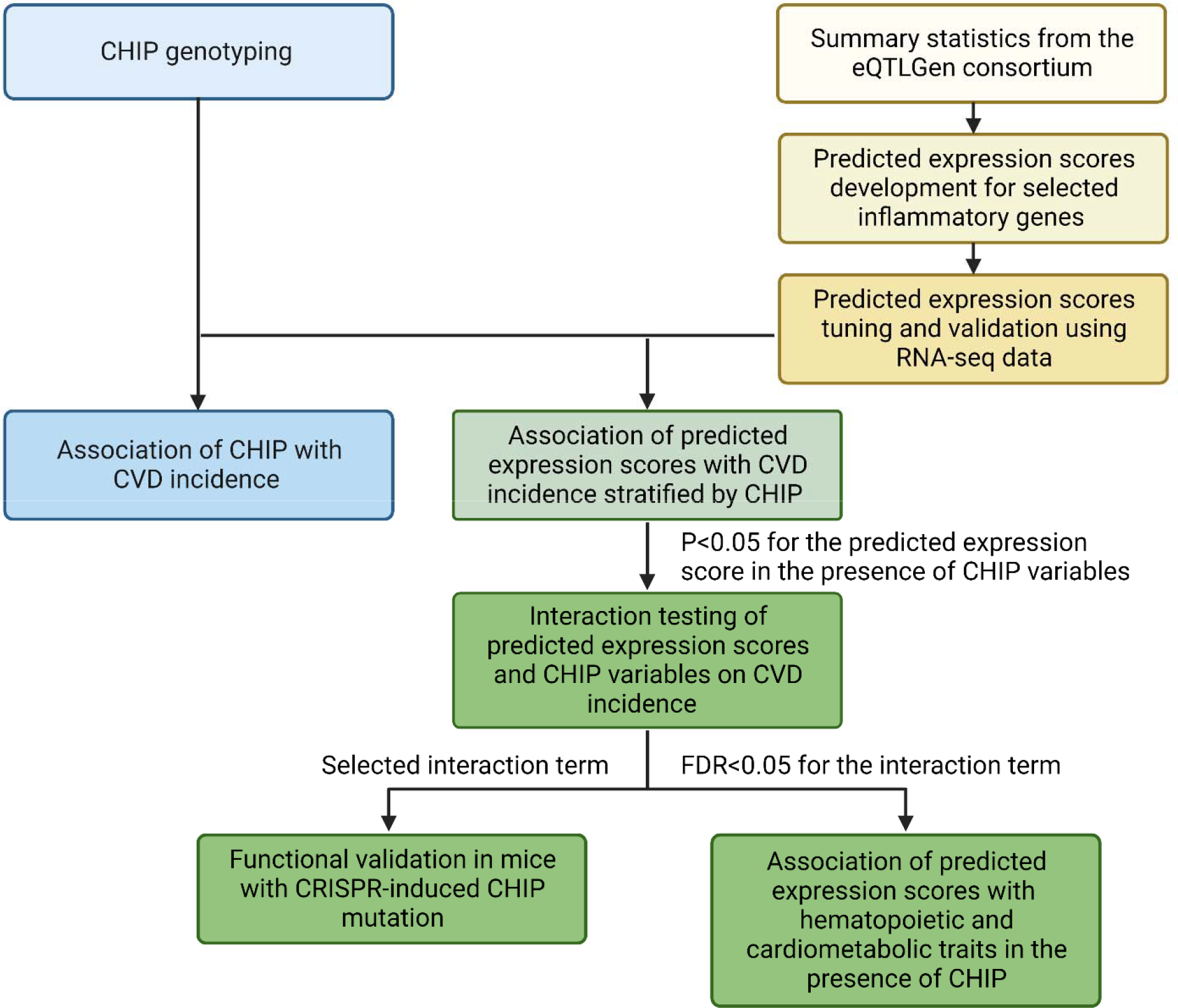
Study schematics. CHIP was identified using whole exome sequencing data of blood DNA. Predicted expression score for inflammatory genes was developed based on *cis*-expression quantitative trait locus results and validated using measured RNA-sequencing data, and then examined whether they modified CHIP-associated CVD risk. Predicted expression scores that significantly modify CHIP-associated CVD risk were further validated in mice model and evaluated for their associations with hematopoietic and cardiometabolic traits. CHIP: clonal hematopoiesis of indeterminate potential; CVD: cardiovascular disease; FDR: false discovery rate; HR: hazard ratio.

**Table 1.**
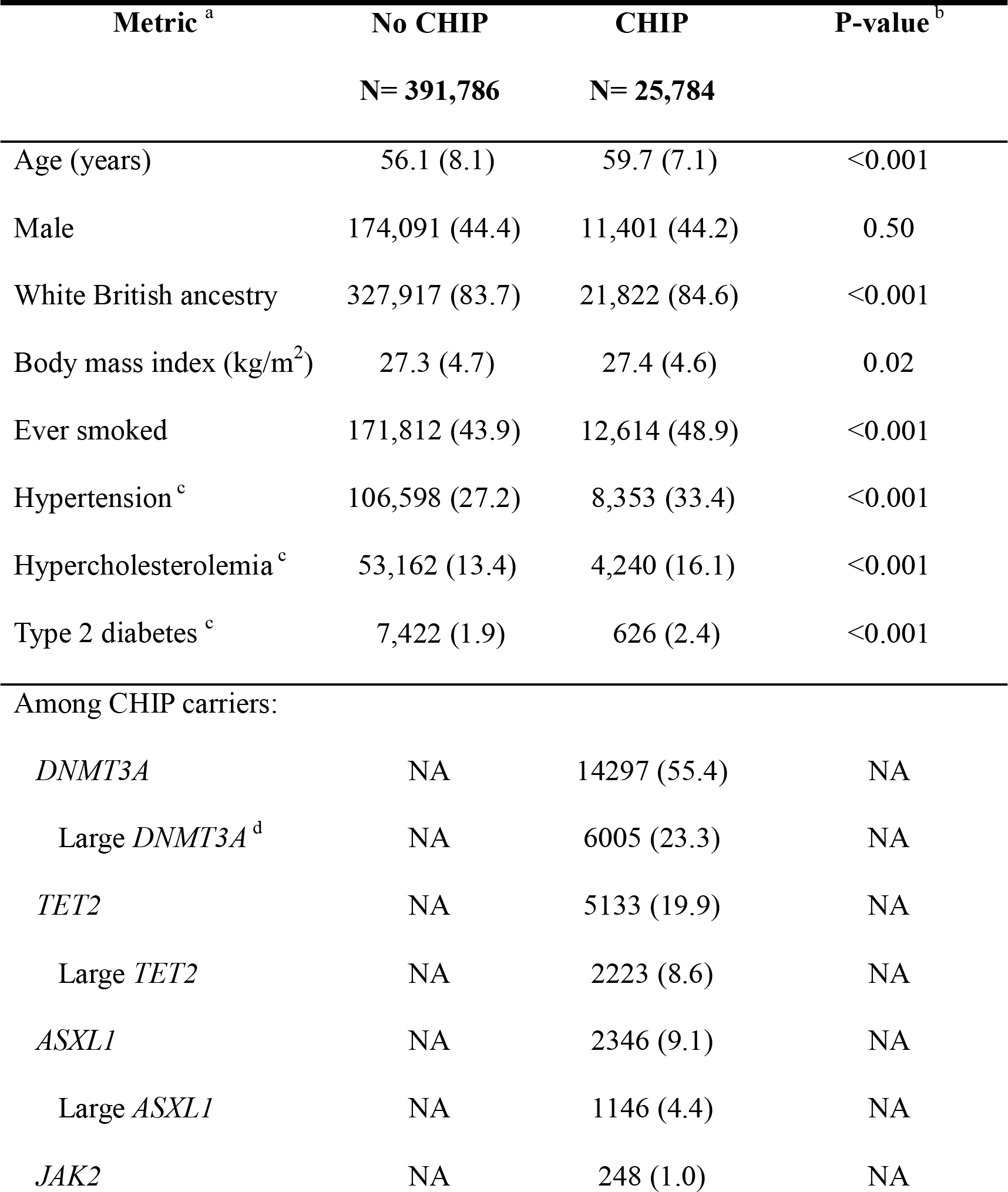
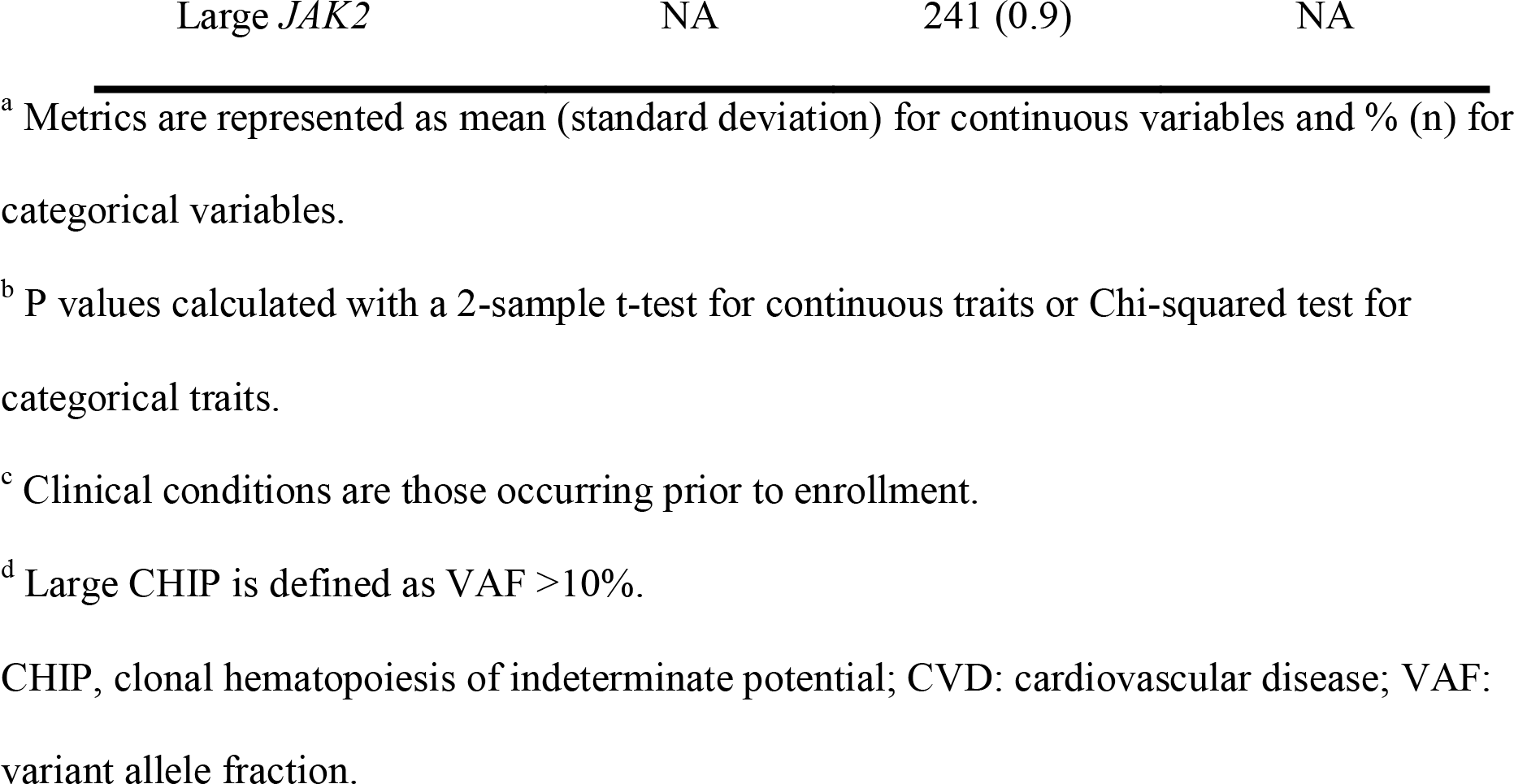
Characteristics of the study population in the UK Biobank (N= 417,570). Study population was restricted to unrelated individuals in the UK Biobank who had exome sequencing data and were free of hematological cancers and composite CVD events at baseline with unrelatedness defined as less than 3rd-degree relatedness.

| Metric <sup>a</sup> | No CHIP<br>N= 391,786 | CHIP<br>N= 25,784 | P-value <sup>b</sup> |
| --- | --- | --- | --- |
| Age (years) | 56.1 (8.1) | 59.7 (7.1) | <0.001 |
| Male | 174,091 (44.4) | 11,401 (44.2) | 0.50 |
| White British ancestry | 327,917 (83.7) | 21,822 (84.6) | <0.001 |
| Body mass index (kg/m<sup>2</sup>) | 27.3 (4.7) | 27.4 (4.6) | 0.02 |
| Ever smoked | 171,812 (43.9) | 12,614 (48.9) | <0.001 |
| Hypertension <sup>c</sup> | 106,598 (27.2) | 8,353 (33.4) | <0.001 |
| Hypercholesterolemia <sup>c</sup> | 53,162 (13.4) | 4,240 (16.1) | <0.001 |
| Type 2 diabetes <sup>c</sup> | 7,422 (1.9) | 626 (2.4) | <0.001 |
| Among CHIP carriers: |  |  |  |
| *DNMT3A* | NA | 14297 (55.4) | NA |
| Large *DNMT3A* <sup>d</sup> | NA | 6005 (23.3) | NA |
| *TET2* | NA | 5133 (19.9) | NA |
| Large *TET2* | NA | 2223 (8.6) | NA |
| *ASXL1* | NA | 2346 (9.1) | NA |
| Large *ASXL1* | NA | 1146 (4.4) | NA |
| *JAK2* | NA | 248 (1.0) | NA |

<sup>a</sup> Metrics are represented as mean (standard deviation) for continuous variables and % (n) for categorical variables.
<sup>b</sup> P values calculated with a 2-sample t-test for continuous traits or Chi-squared test for categorical traits.
<sup>c</sup> Clinical conditions are those occurring prior to enrollment.
<sup>d</sup> Large CHIP is defined as VAF >10%.
CHIP, clonal hematopoiesis of indeterminate potential; CVD: cardiovascular disease; VAF: variant allele fraction.

### Associations between CHIP mutations and incident CVD

During the 11.0-year median follow-up, 44962 (10.6%) incident CVD events (a composite of myocardial infarction, coronary artery disease or revascularization, stroke, or death) (7) were observed. The presence of any CHIP associated with increased CVD event risk independent of potential confounders (age, sex, white British ancestry, body mass index [BMI] at the time of enrollment, ever-smoker status, diagnoses of type 2 diabetes mellitus at the time of enrollment, and the first ten principal components of genetic ancestry) with a composite effect of hazard ratio (HR) 1.18 (95% confidence interval [CI]: 1.14-1.22, *P*: 1.5×10^-21^). Among the top CHIP genes, CVD effects varied by genes with *JAK2* 2.81-fold (95% CI: 2.25-3.51, P-value: 8.5×10^-20^), *ASXL1* 1.41-fold (95% CI: 1.29-1.54, *P*: 3.5×10^-14^), *TET2* 1.11-fold (95% CI: 1.03-1.19, *P*: 4.5×10^-3^), and *DNMT3A* 1.06-fold (95% CI: 1.01-1.11, *P*: 0.01). In addition, other CHIP genes also showed significant associations with CVD incidence with *SRSF2* 2.6-fold (95% CI: 2.18-3.09, P-value: 6.8×10^-27^), *SF3B1* 1.47-fold (95% CI: 1.14-1.89, *P*: 2.9×10^-3^), *TP53* 1.43-fold (95% CI: 1.18-1.72, *P*: 2.2×10^-4^), and *PPM1D* 1.39-fold (95% CI: 1.18-1.64, *P*: 7.6×10^-5^). Large clones generally demonstrated larger effects, with large CHIP associated with 1.29-fold (95% CI: 1.24-1.35, P: 8.6x10^-29^) incident CVD risk.

(**Table 2**) (16). Sensitivity analyses restricting the outcome to coronary artery disease (CAD) alone yielded attenuated results in the same directions (**Supplemental Table 1**)

**Table 2.** Associations between CHIP mutation and incidence of CVD event. CVD event outcome is defined as a composite of myocardial infarction, coronary artery disease or revascularization, stroke, or death. b Models were adjusted for age at the time of enrollment, sex, white British ancestry, body mass index, diagnoses of type 2 diabetes mellitus at the time of enrollment, diagnoses of hypertension at the time of enrollment, ever-smoker status, and the first ten principal components of genetic ancestry. Participants with prevalent hematological cancers or CVD were removed from the analyses.

|  | Presence of CHIP |  | Presence of large CHIP<sup>a</sup> |  |
| --- | --- | --- | --- | --- |
|  | HR (95% CI) | P-value | HR (95% CI) | P-value |
| **CHIP** | 1.18 (1.14, 1.22) | $1.5 \times 10^{-21}$ | 1.29 (1.24, 1.35) | $8.6 \times 10^{-29}$ |
| *DNMT3A* | 1.06 (1.01, 1.11) | 0.01 | 1.13 (1.06, 1.21) | $3.9 \times 10^{-4}$ |
| *TET2* | 1.11 (1.03, 1.19) | $4.5 \times 10^{-3}$ | 1.28 (1.16, 1.41) | $5.8 \times 10^{-7}$ |
| *ASXL1* | 1.41 (1.29, 1.54) | $3.5 \times 10^{-14}$ | 1.52 (1.35, 1.71) | $4.1 \times 10^{-12}$ |
| *JAK2* | 2.81 (2.25, 3.51) | $8.5 \times 10^{-20}$ | 2.80 (2.23, 3.50) | $3.3 \times 10^{-19}$ |
| *PPM1D* | 1.39 (1.18, 1.64) | $7.6 \times 10^{-5}$ | 1.46 (1.13, 1.88) | $3.9 \times 10^{-3}$ |
| *TP53* | 1.43 (1.18, 1.72) | $2.2 \times 10^{-4}$ | 2.04 (1.57, 2.65) | $1.0 \times 10^{-7}$ |
| *SRSF2* | 2.60 (2.18, 3.09) | $6.8 \times 10^{-27}$ | 3.32 (2.72, 4.04) | $1.1 \times 10^{-32}$ |
| *SF3B1* | 1.47 (1.14, 1.89) | $2.9 \times 10^{-3}$ | 1.55 (1.16, 2.06) | $2.9 \times 10^{-3}$ |

<sup>a</sup> Large CHIP is defined as VAF >10%.

### Predicted expression of inflammatory genes

We expanded the examination for CHIP modifiers through two dimensions. (1) In addition to a composite of all CHIP mutations at any driver genes, we examined the most commonly mutated CHIP genes individually (6), such as *DNMT3A*, *TET2*, *ASXL1,* and *JAK2*. (2) In addition to *IL6R*, we generated predicted expression levels of all other inflammatory genes that are implicated in or closely related to the NLRP3 and AIM2 inflammasome pathways, including *NLRP3*, *IL1B*, *IFNG*, *IL18*, *CARD8, CASP1, CASP5, DHX33, IFNGR1, IFNGR2, IL1R1, IL1R2, IL1RAP, IL6, IL6ST, IL10, IL18BP, IL18R1, IL18RAP, IRF1, JAK1, JAK2, JAK3, NEK7, NLRC4, SOCS, STAT1, STAT3, STAT4, STAT5A, STAT6, TNF,* and *TYK2* (**Methods** and **Supplemental Figure 1**).

We developed predicted expression scores based on summary statistics of the whole blood or peripheral blood mononuclear cells (PBMC) cis-expression quantitative trait locus (eQTL) results for the corresponding genes from the eQTLGen Consortium (20). For each selected gene, we used both the pruning and thresholding method (P+T) (21) and the polygenic risk score-continuous shrinkage (PRS-CS) method (22) to generate a series of candidate scores for participants with European ancestry (EA) and non-European ancestry separately; they were then tuned using non-overlapping individual-level RNA-sequencing data from the Framingham Study (FHS; whole blood) and Multi-Ethnic Study of Atherosclerosis (MESA; PBMC) (23, 24). The final predicted expression score of each gene was selected based on the proportion of the variance (R^2^) of experimentally measured expression levels that can be explained by the candidate scores (**Methods**). For most genes, the P+T method generated a better score performance than PRS-CS (**Supplemental Table 2**). For this analysis, we kept genes whose selected best-performing predicted expression scores had R^2^ >1% among EA participants, resulting in scores for 26 (of 35 total evaluated) genes. The predicted expression scores explained a median of 3.5% [interquartile range 1.8%-6.3%] of the adjusted variance of corresponding gene expression levels among EA participants. The score for *IL18RAP* explained the largest proportion of phenotypic variance (34.7%), and that for *IL1B* explaining the least (1.05%) among analyzed genes with R^2^>1% (**Figure 2 and Supplemental Table 2**).

**Figure 2.**
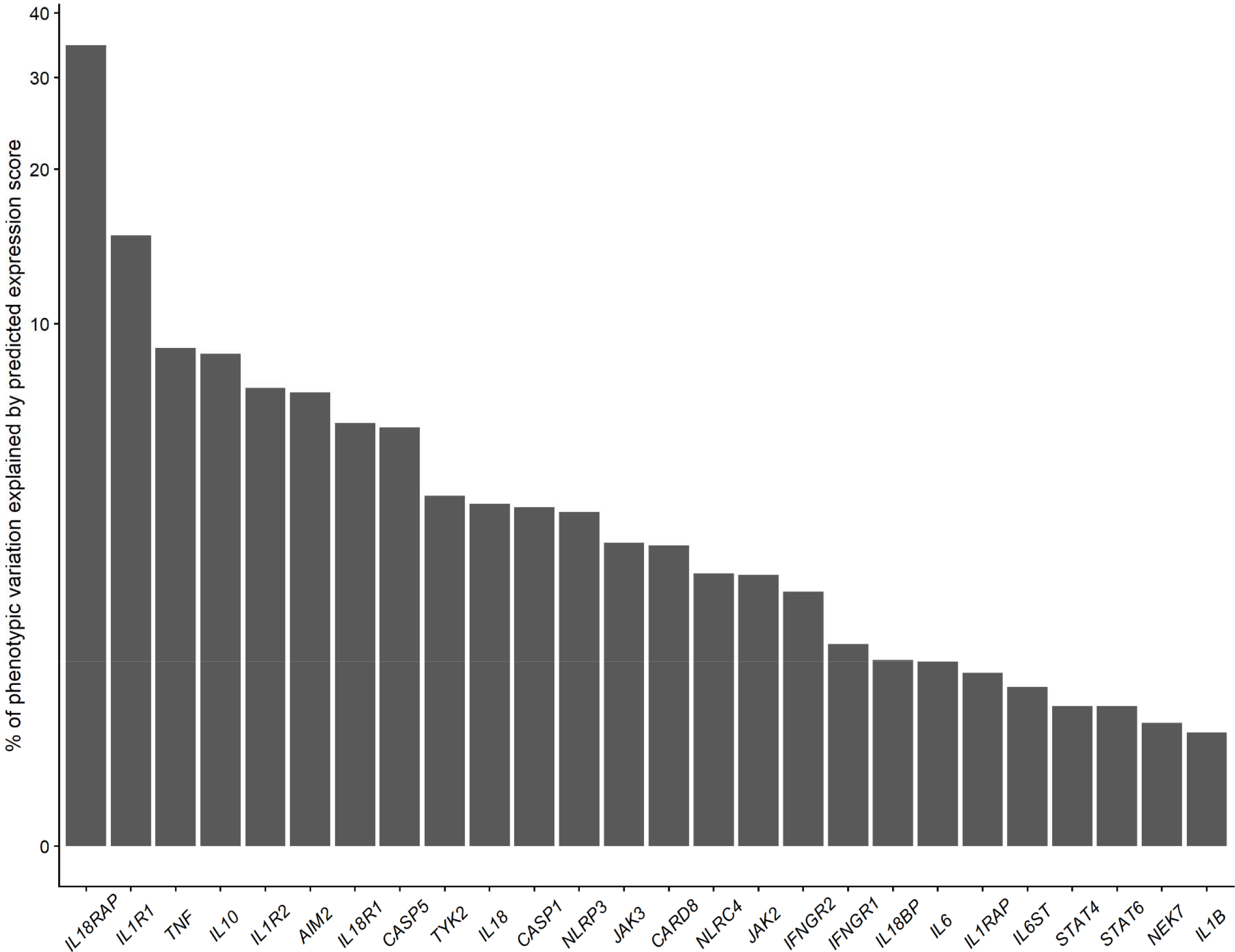
Proportion of the variance of experimentally measured expression levels that can be explained by predicted expression scores for inflammatory genes among participants with European ancestry. Inflammatory genes were identified through canonical pathways and protein- protein interactions based on STRING (https://string-db.org/). Predicted expression scores of examined genes were calculated by applying either P+T or PRS-CS methods to the summary statistics of the eQTL for those genes from the eQTLGen consortium (https://www.eqtlgen.org/) and validated using experimental measured RNA-sequencing data in the Multi-Ethnic Study of Atherosclerosis (peripheral blood mononuclear cells [PBMC]) and Framingham Heart Study (whole blood). Since the eQTL source data was from either PBMC or whole blood, we report the largest R^2^ of the measured transcriptome levels in either FHS or MESA. eQTL: expression quantitative trait loci; PRS-CS: polygenic risk score-continuous shrinkage; P+T: pruning and thresholding

### Modification of CHIP-associated CVD risk by predicted expression of inflammatory genes

We observed significant associations between predicted expression scores of several inflammatory genes and incident CVD risk with the presence of CHIP or specific CHIP gene(s) (collectively called CHIP variables), while the corresponding associations for those without CHIP were all non-significant. For predicted expression scores that were significantly associated with incident CVD risk at *P*<0.05 level only in the presence of CHIP variable(s), we carried forward to evaluate the interactions between those scores and the corresponding CHIP variables (N=9 pairs) on the primary CVD outcome (**Figures 3 and 4**).

**Figure 3.**
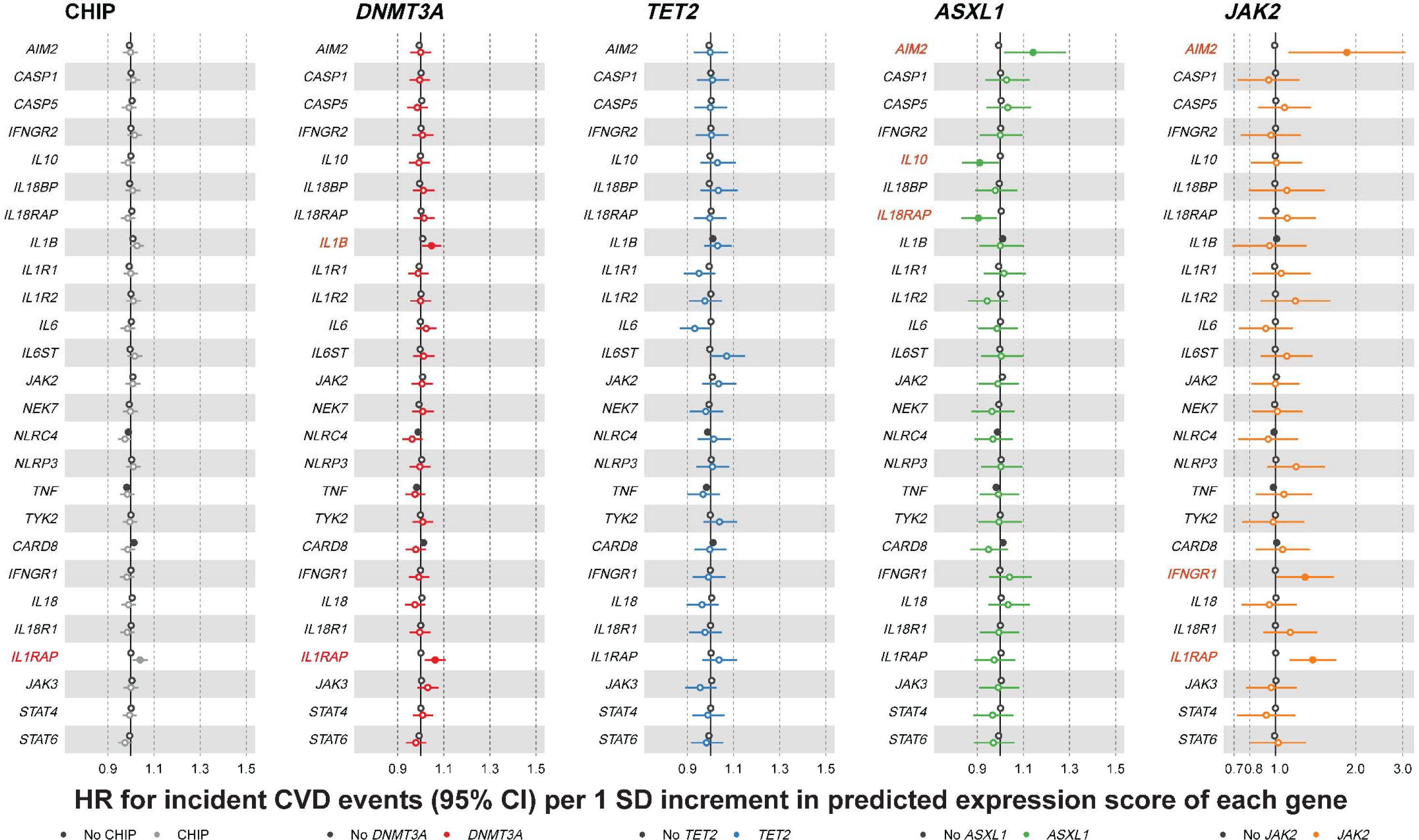
HR of predicted expression scores of inflammatory genes on CVD event incidence stratified by CHIP mutation status. Inflammatory genes were identified through canonical pathways and protein-protein interactions based on STRING (https://string-db.org/). Predicted expression scores of examined genes were calculated by applying either P+T or PRS-CS methods to the summary statistics of the eQTL for those genes from the eQTLGen consortium (https://www.eqtlgen.org/) and validated using experimental measured RNA-sequencing data in the Multi-Ethnic Study of Atherosclerosis (peripheral blood mononuclear cells) and Framingham Heart Study (whole blood). CVD event outcome is defined as a composite of myocardial infarction, coronary artery disease or revascularization, stroke, or death. Black color indicates the absence of CHIP mutations, and non-black colors indicate the presence of CHIP mutations. A solid circle indicates a significant association at *P*<0.05 level. Red colored gene name indicates significant association between corresponding expression score and CVD outcome in the presence of CHIP mutation at *P*<0.05 level. CHIP, clonal hematopoiesis of indeterminate potential; CVD, cardiovascular disease; eQTL: expression quantitative trait loci; HR: hazard ratio; PRS-CS: polygenic risk score-continuous shrinkage; P+T: pruning and thresholding

**Figure 4.**
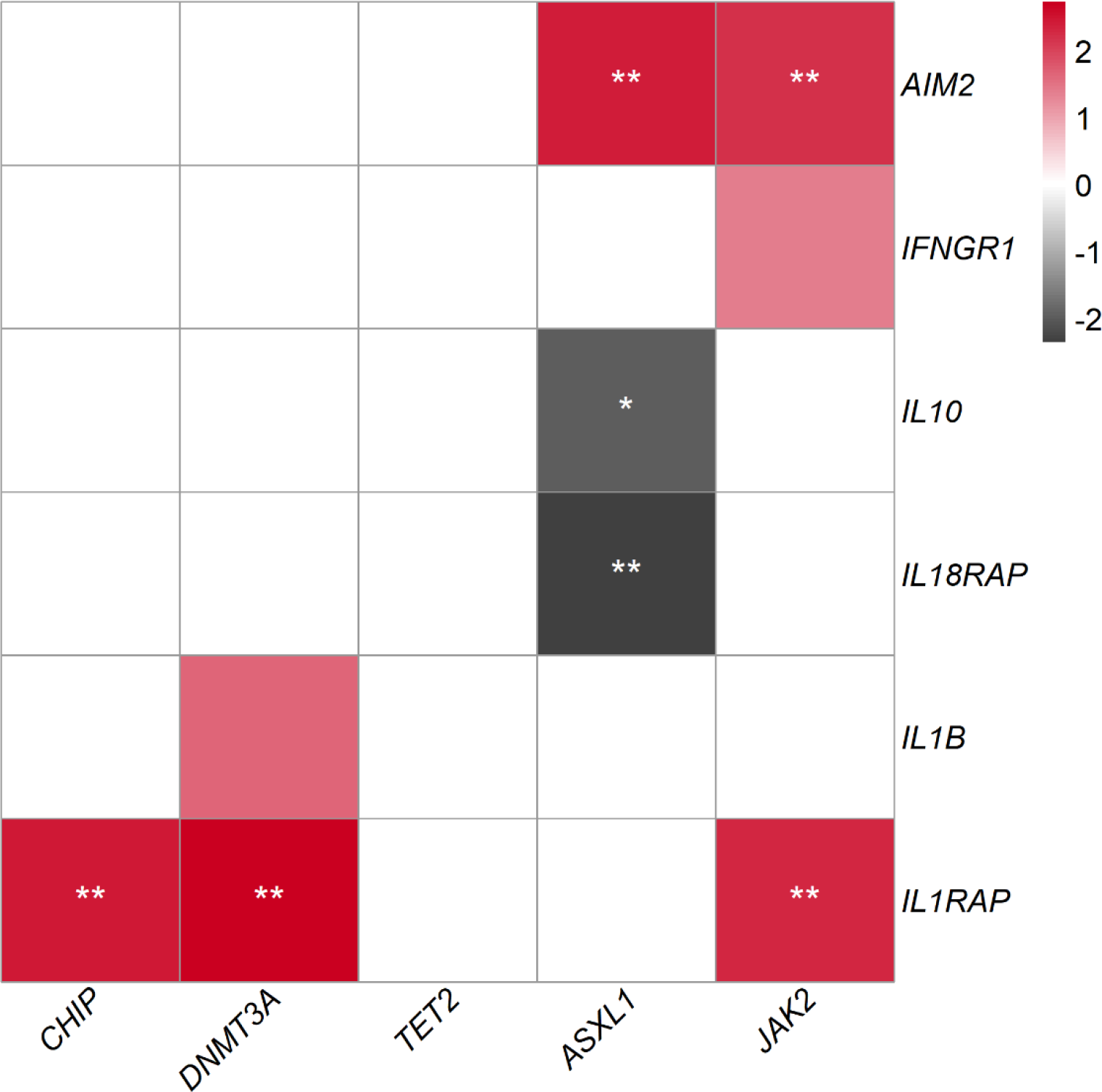
Heatmap for the Z-scores of interactions between CHIP mutations and predicted expression scores of inflammatory genes on CVD event incidence. Only predicted expression scores significantly associated with CVD event incidence among participants with CHIP mutations were examined for their interactions in this step. Inflammatory genes were identified through canonical pathways and protein-protein interactions based on STRING (https://string-db.org/). Predicted expression scores of examined genes were calculated by applying either P+T or PRS-CS methods to the summary statistics of the eQTL for those genes from the eQTLGen consortium (https://www.eqtlgen.org/). CVD event outcome is defined as a composite of myocardial infarction, coronary artery disease or revascularization, stroke, or death. Black color indicates a negative Z-score, and red indicates a positive Z-score. Two white stars indicate statistical significance at an FDR=0.05 level; a single white star indicates statistical significance at an FDR=0.1 level. The darker the color, the stronger the effects. CHIP, clonal hematopoiesis of indeterminate potential; CVD, cardiovascular disease; eQTL: expression quantitative trait loci; FDR: false discovery rate; PRS-CS: polygenic risk score-continuous shrinkage; P+T: pruning and thresholding

Regarding specific modification pairs, first, we found evidence supporting recent murine findings (15) in humans by observing that a genetic predisposition to higher *AIM2* expression was associated with amplified risk for incident CVD for those with *JAK2* CHIP. One standard deviation (SD) increase in predicted expression score for *AIM2* was associated with an almost 2- fold increased risk in CVD incidence (HR: 1.85, 95% CI: 1.12-3.07, *P*: 0.02) among participants with *JAK2* mutations. In contrast, the predicted expression score for *AIM2* was not associated with incident CVD event risk in those without *JAK2* mutations (HR, 0.99; 95% CI: 0.98–1.00; *P*: 0.16), which was significantly different for those with *JAK2* CHIP (false discovery rate [FDR] for interaction: 0.04). Mice expressing *Jak2*^VF^ in bone marrow had a 2-fold increase in atherosclerotic lesion development which was markedly reduced by genetic ablation of *Aim2* in mutant cells (15). Moreover, the CVD risk associated with *JAK2^VF^* CHIP was augmented by higher predicted expression of *IFNGR1*. Interferon-γ increased *Aim2* expression in *Jak2^VF^* BMDMs and AIM2 levels were increased in plaques of *Jak2^VF^* CHIP mice (15). These findings support the translational relevance of murine models of *Jak2^VF^* CHIP.

Second, we observed modification effects of the predicted expression level of *IL1RAP* on incident CVD risk associated with any CHIP, *DNMT3A*, and *JAK2* CHIP mutations. *IL1RAP* encodes IL-1 receptor accessory protein (IL-1RAP), a coreceptor involved in several inflammatory signaling pathways and the lack of which completely abrogates cellular response to IL-1 (25–28). For one standard deviation (SD) increase in the predicted expression score for *IL1RAP*, HRs (95% CI) were 1.04 (1.01, 1.07) in the presence of any CHIP mutations, 1.06 (1.02, 1.11) in the presence of *DNMT3A* mutation, and 1.38 (1.13, 1.69) in the presence of *JAK2* mutations, in contrast with HRs (95% CI) of 1.00 (0.99, 1.01), 1.00 (0.99, 1.01), and 1.00 (0.99, 1.01) among participants without these mutations (FDR for interaction: 0.04, 0.04, and 0.04, respectively). While the relationship for *TET2* was directionally consistent, no significant association was observed. This result implicates IL1RAP as a potentially key IL-1β/IL-6 pathway-related molecule for CHIP-associated CVD risk across genes (7, 11).

Third, we identified novel potential modification effects by the predicted expression of *AIM2* and *IL10* on *ASXL1*-associated CVD risk. In addition to it *AIM2*’s aforementioned interaction with *JAK2* on CVD risk, *AIM2* predicted expression showed a similar modification effect on the *ASXL1*-associated CVD disease risk (*ASXL1* mutation present: HR: 1.14, 95% CI: 1.02-1.28; *ASXL1* mutation absent: HR: 0.99, 95% CI: 0.98-1.00; FDR for interaction: 0.04). Similar effects were not observed for *DNMT3A* or *TET2*-associated CVD. *IL10* is expressed in atherosclerotic plaques, and its encoded protein, IL-10, is an anti-inflammatory cytokine that inhibits many cellular processes that advance human atherosclerosis (29–37). The protective effect of IL-10 is pronounced in the presence of *ASXL1* mutation, with its predicted expression score associated with a significantly decreased risk of incident CVD (HR, 0.91; 95% CI: 0.83– 0.99; *P*: 0.04) in the presence of *ASXL1* mutation but a null effect (HR, 1.00; 95% CI: 0.99– 1.01; *P*: 0.91) when it is absent (FDR for interaction: 0.06). Another molecule implicated was *IL18RAP* which encodes IL-18 receptor accessory protein (IL-18RAP). IL-18RAP enhances the IL-18-binding activity of the IL-18 receptor and plays a role in signaling by the inflammatory cytokine IL-18 (38). However, we observed attenuated CVD risk associated with the predicted expression score of *IL18RAP* among participants with *ASXL1* mutation (HR: 0.90, 95% CI: 0.83- 0.98, *P*: 0.02) but not those without (HR: 1.00, 95% CI: 0.99-1.01, *P*: 0.41; FDR for interaction: 0.04). These results are shown in **Figure 3**, **Figure 4, Supplemental Table 3,** and **Supplemental Table 4**. These identified inflammatory expression scores that modify CHIP variable-associated CVD risk were not associated with the corresponding CHIP variable, with *JAK2* gene expression and *JAK2* CHIP mutation (FDR=6.1×10^-6^) as the exception.

### AIM2 inflammasome activation in macrophages harboring Asxl1 mutations

Our findings indicated that the predicted expression score of *AIM2* was associated with an increased risk of CVD events in patients with *JAK2* and *ASXL1* clonal hematopoiesis (**Figure 3 and Figure 4**). While AIM2 inflammasome activation has been linked to *JAK2* clonal hematopoiesis (15, 39), the AIM2 inflammasome has not previously been associated with *ASXL1*. To understand if *Asxl1* mutations promote AIM2 inflammasome activation, we introduced truncation mutations into hematopoietic stem and progenitor cells (HSPCs) in exon 12 of *Asxl1* using CRISPR (**Figure 5A**). Bone marrow-derived macrophages (BMDMs) from mice with CRISPR guides (Control) or *Asxl1* mutations (*Asxl1-G623*\*) showed no genotype-dependent alteration in NLRP3 inflammasome activation when challenged with LPS and ATP (**Figure 5B**). In contrast, *Asxl1* mutant macrophages demonstrated a selective increase in AIM2 inflammasome activation when treated with the double-stranded DNA fragments (pdAdT) (**Figure 5B**). Consistent with increased inflammasome activation, *Asxl1* mutant macrophages had increased LPS-induced *Il1b* production without altered *Casp1* or *Il1rap* expression (**Figure 5C-E**). LPS-induced *Nlrp3* expression was reduced in *Asxl1* mutant macrophages (**Figure 5F**), which may explain why we did not observe increased NLRP3 activation even in the presence of increased *Il1b*. *Aim2* expression was unchanged in *Asxl1* mutant macrophages (**Figure 5G**). Since the AIM2 inflammasome may be activated in response to DNA damage, we measured pγH2AX, a marker of nuclear DNA damage and double-strand break formation (40), and found a significant increase in pγH2AX in *Asxl1* mutant cells (**Figure 5H-I**). These observations suggest that *Asxl1* mutant macrophages have increased *Il1b* expression and increased DNA damage that together lead to increased AIM2 inflammasome activation.

**Figure 5.**
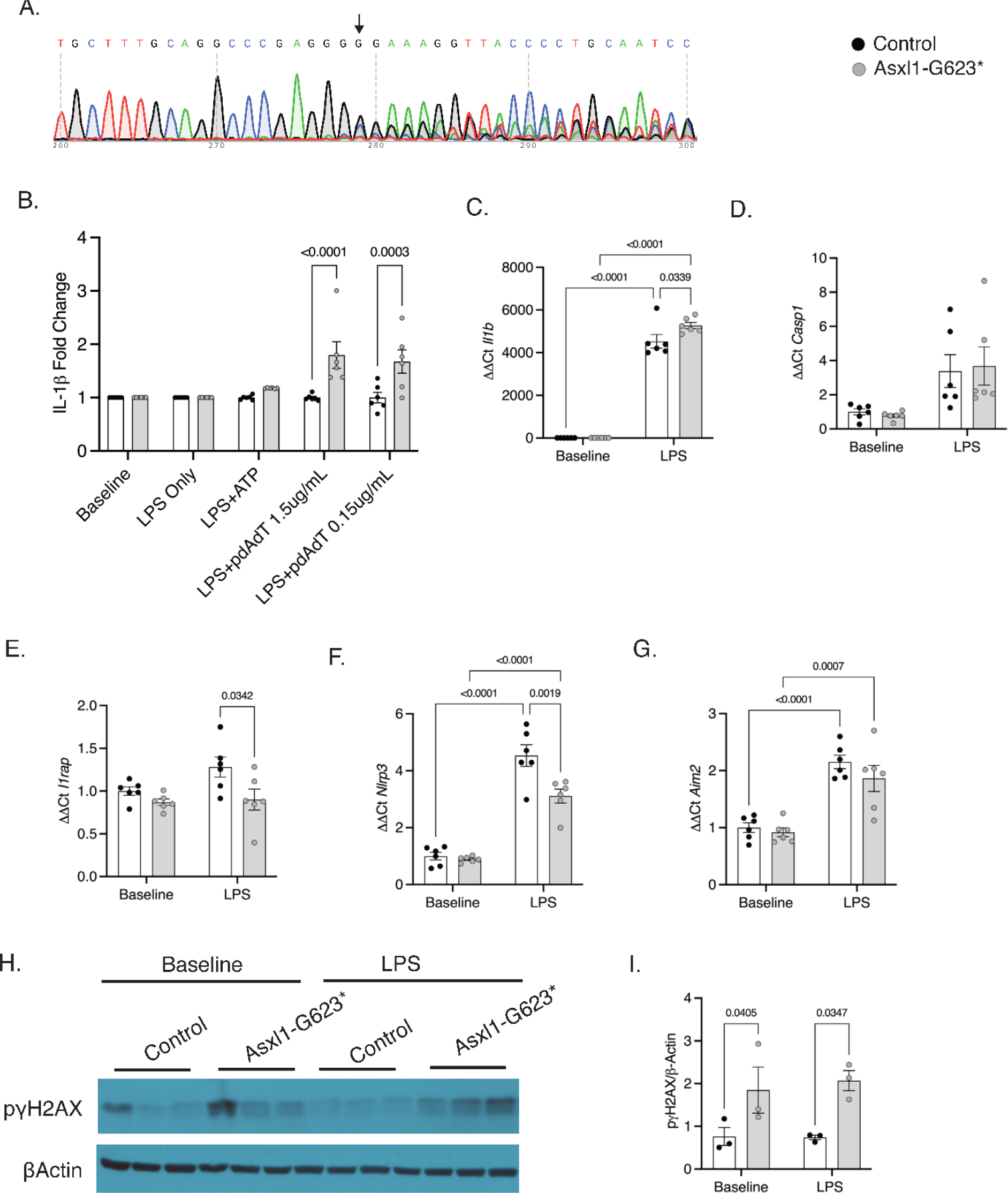
Inflammasome activation in bone marrow derived macrophages harboring *Asxl1* mutations. Bone marrow derived macrophages (BMDMs) were harvested from mice harboring either a mixture of wild- type control (Nmt4) or Asxl1 mutated bone marrow (*Asxl1-G623*\*) with WT bone marrow. A. Sanger Sequencing of Cas9 transgenic murine fibroblast transfected with lentiviruses containing Asxl1-guides targeting exon 12, arrow indicates target site. B. Inflammasome activation marked by IL-1β in supernatant of BMDM primed with LPS then ATP was used for stimulating NLRP3 inflammasome or poly (deoxyadenylic- deoxythymidylic) (pdAdT) for AIM2 inflammasome activation; Data are fold change. C-G. qPCR analysis of BMDMs at baseline or following 6-hour stimulation with 20ng/ml lPS. H. Western blot analysis of BMDMs at baseline or following 6-hour stimulation with 20ng/mL LPS. I. Densiometric quantification of western blot. Data are mean±Standard error of the mean. Two-way ANOVA followed by Tukey post hoc test B-G & I.

### Asxl1 mutant macrophages have pro- and anti-inflammatory characteristics

Although AIM2 inflammasome activation has been shown to be sufficient to promote atherosclerosis in *Jak2* clonal hematopoiesis (15), our findings suggest other pathways may also contribute to *ASXL1*- mediated CVD risk (**Figure 3 and Figure 4**). Therefore, we examined inflammatory mediators secreted by BMDMs under baseline and LPS-stimulated conditions. In response to LPS, *Asxl1* mutant macrophages had no change in *Il6* expression; however, IL-6 secretion was increased (Figure **6A & 6B**), which is consistent with elevated IL-6 in serum from patients with *ASXL1* clonal hematopoiesis (41). Interestingly, *Tnfa* expression and secretion were both reduced in *Asxl1* mutant macrophages (**Figure 6C-D**), while we did not see a similar suppression of other LPS-sensitive genes such as *Il1b*, *Il6*, *Il1a*, *Ccl3*, or *Tgfb* (**Figure 5C**, **Figure 6A, and Figure 6E-F**). These observations suggest that LPS-induced inflammatory signaling is largely intact in *Asxl1* mutant macrophages, some components may be disrupted potentially due to *Asxl1*- mediated changes in chromatin accessibility. We found that the predicted expression of *IL10* may play a protective role in *ASXL1*-mediated CVD risk in humans (**Figure 3 and Figure 4**), and IL-10 is also a potent inhibitor of tumor necrosis factor-alpha (TNFα). Therefore, we examined if IL-10 was dysregulated in the presence of *Asxl1* mutations. We observed that stimulation with LPS increased the anti-inflammatory mediator *Il10* more than two-fold in *Asxl1* mutant BMDMs compared to control, and resulted in a similar increase in secreted IL-10 (**Figure 6H-I**), paralleled by an increase in the *Il10* target gene *Socs3* (**Figure 6J**). *Socs3* was also found to be increased in *Asxl1* mutant zebrafish (42). Thus, our population genetic data identified the predicted expression of *IL10* as a potential suppressor of *ASXL1*-mediated CVD disease, which is supported by functional studies suggesting that IL-10 levels and signaling are increased in *Asxl1* mutant macrophages and may play an important role in inflammation regulation.

**Figure 6.**
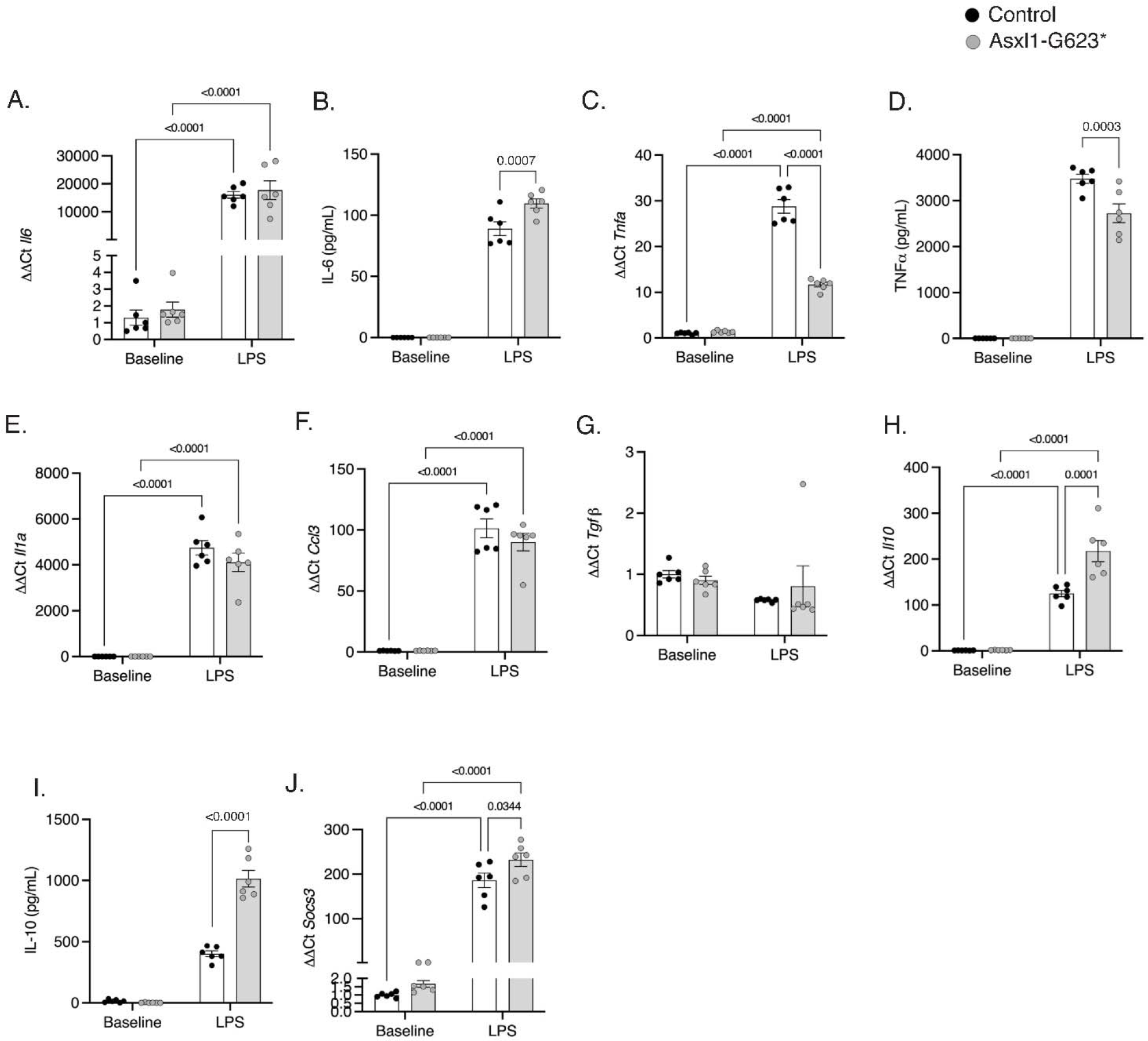
*Asxl1* mutant macrophages have pro- and anti-inflammatory characteristics. Bone marrow derived macrophages (BMDMs) were untreated (baseline) or treated with 20ng/mL LPS for 6 hours. A. qPCR analysis. B. ELISA quantification of protein in culture media. C. qPCR analysis. D. ELISA quantification of protein in culture media. E-H. qPCR analysis. I. ELISA quantification of protein in culture media. J. qPCR analysis. Data are mean±Standard error of the mean. Two-way ANOVA followed by Tukey post hoc test A-J.

### Asxl1 mutations and atherosclerosis

To determine the impact of *Asxl1* on atherosclerosis, we attempted to model *Asxl1* clonal hematopoiesis by transplanting CD45.2^+^Cas9^+^ transgenic LT- HSCs infected with control (non-targeting guide RNAs) or *Asxl1-G623\** guide RNAs mixed with CD45.1^+^WT cells into lethally irradiated *Ldlr^-/-^*mice. Mice were then placed on a Western- type diet (WTD) to induce hypercholesteremia (**Figure 7A**). *Asxl1* mutations did not alter leukocyte counts in blood or spleen weight (**Figure 7B-F**). Evaluation of Asxl1 mutant burden in blood cells indicated that *Asxl1* mutant cells made up only ∼15% of lymphocytes, ∼5% of neutrophils, and ∼2% of blood monocytes by the end of the study (**Figure 7G-I**), indicating a very low mutation burden in these animals. Histological analysis of aortic root lesions indicated no change in the lesion area or necrotic core area (**Figure 7J-L**). Our current observations are consistent with previous reports showing impaired initial HSC proliferation and clonal expansion in *Asxl1* CHIP mice, and suggest a much longer follow-up time (> one year) may be needed to promote atherosclerosis development in the *Asxl1* mice model (43).

**Figure 7.**
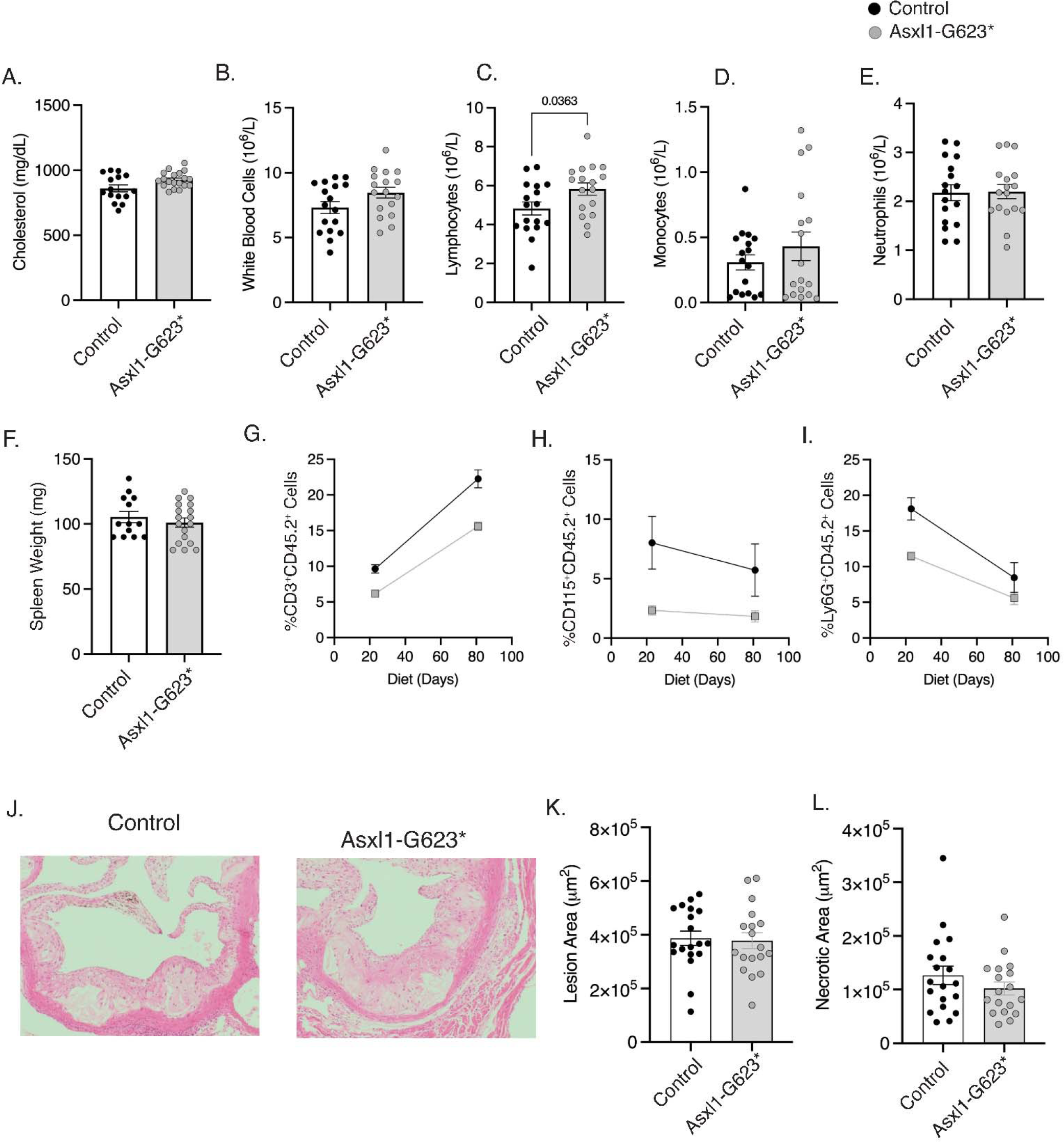
*Asxl1* mutations and atherosclerosis. Mice transplanted with chimeric mixtures of bone with non- targeting guide RNAs (Control) and *Asxl1-G623*\* guides. A. Terminal serum cholesterol. Complete blood cell counts at the end of western type diet feeding of B. White blood cells, C. Lymphocytes, D. Monocytes, and E. Neutrophils. F. Spleen weight. G. %CD45.2^+^ mutated cells in blood G. Lymphocytes, H. Monocytes, and I. Neutrophils. J. Representative images of Hemotoxin and Eosin (H&E) stained aortic root lesions and K. quantification of lesion area and L. necrotic core area. Data are mean±Standard error of the mean. Students t- test A-F, K, & L. Two-way ANOVA followed by Tukey post hoc test G-I.

### Associations with hematopoietic traits and cardiometabolic biomarkers

For the eight CHIP mutation-predicted expression score pairs that showed significant modification effects on CVD incidence, we examined the associations between those predicted expression scores with 31 hematopoietic traits and five common cardiometabolic biomarkers among participants with the corresponding CHIP mutations. After accounting for multiple hypothesis testing [N=248 (8*31) for hematopoietic traits and N=40 (8*5) for cardiometabolic biomarkers], we did not observe any significant associations passing the FDR=0.05 threshold. The suggestive nominal associations were observed between the predicted expression score of *IL18RAP* and reduced eosinophil count and eosinophil percentage among individuals with *ASXL1* mutations (P=0.002 and 0.003, respectively). This is in line with previous analysis of cap analysis of gene expression (CAGE) sequencing data showed that *IL18RAP* is highly expressed in eosinophils, neutrophils, and natural killer (NK) cells (44) **(Supplemental Figure 2**, **Supplemental Table 5**, and Supplemental Table 6).

## Discussion

Leveraging validated human genetic instruments, we show that specific inflammatory genes may influence incident CVD risk in a manner that is specific to the presence of mutations in key CHIP genes. Our observations are consistent with the notions that reduced *AIM2* expression could specifically mitigate *JAK2* mutation-associated CVD risk and with IL1RAP as a key molecule for CHIP-associated CVD risk across multiple CHIP genes; these findings agree with prior murine studies. Furthermore, we discovered potential modification by *AIM2* expression on *ASXL1*-associated CVD risk in humans with corroboration in CRISPR-induced *Asxl1* mutated murine BMDMs. Our observations provide human genetic and pre-clinical support toward new precision medicine paradigms for CVD meriting assessment in prospective studies.

Our study has three key implications. First, our findings further show that CVD prognosis and mechanism are distinct by implicated CHIP gene. Prior studies showed that NLRP3 inflammasome inhibition mitigates the heightened atherogenesis observed in *Tet2* chimeric atherogenic mice compared to atherogenic mice wild-type for *Tet2* (11). Correspondingly, a common disruptive coding variant in *IL6R* (a downstream mediator of NLRP3) *TET2* or *DNMT3A* CHIP among humans (7, 45). A *post hoc* exploratory analysis of a completed clinical trial of a monoclonal antibody targeting IL-1B (also a downstream mediator of NLRP3) supports this finding (46). Recently, it was observed that atherogenic mice expressing *Jak2^VF^* displayed a 2-fold increase in atherosclerotic lesion area with increased features of plaque instability that were reduced in the presence of hematopoietic *Aim2* deficiency. The present study used human genetics as instruments and observed similar attenuation effects by genetically-predicted lower expression levels of *AIM2* on *JAK2*-associated CVD risk. These data lend support for AIM2 inflammasome inhibition to address *JAK2*-associated increased CVD risk.

Furthermore, we discovered AIM2’s potential modulatory role for *ASXL1*-associated CVD risk in humans and validated it by demonstrating increased AIM2 inflammasome activation in bone marrow-derived macrophages harboring CRISPR-induced *Asxl1* mutation. In contrast, *Asxl1* mutations did not alter NLRP3 inflammasome activation, which is implicated in *TET2*- associated CAD (11). We further explored the underlying mechanisms. Prior studies showed that *Asxl1* mutant knock-in mice had elevated reactive oxygen species and increased DNA damage (43), and our work further links the induced DNA damage to AIM2 inflammasome activation.

Regarding the proposed mechanism, we note that mutated *ASXL1* forms a complex with BAP1, leading to enhanced histone deubiquitylation activity. Given the well-documented role of BAP1 in the DNA damage response through post-translational modifications of histones (47, 48), it is likely that binding of BAP1 to mutated *ASXL1* may suppress the DNA damage response pathway, causing double-strand DNA breaks to accumulate.

Second, our *Asxl1* mutant macrophage experiments demonstrated both pro- and anti- inflammatory properties, a feature of *Asxl1* that has been previously reported in zebrafish by Avagyan et al. (42). Our study revealed a complex expression profile in *Asxl1* mutant macrophages, potentially linked to alterations in chromatin architecture due to direct histone modifications by *ASXL1* (*49*). Despite noting an increase in IL-6 secretion, our results also demonstrated a decrease in *Tnfa* expression and secretion. Concurrently, we found an increase in *Il10*, a *Tnfa* inhibitor, expression and secretion in *Asxl1* mutant macrophages in murine models.

Concordantly, increased predicted IL10 expression was associated with reduced CVD risk in *ASXL1* CHIP. Together these findings could indicate an important anti-inflammatory role of IL- 10 expression linked to suppression of CVD in *ASXL1* CHIP.

Third, we observed that increased genetic predisposition to *IL1RAP* expression yields increased incident CVD risk for participants with both *DNMT3A* or *JAK2* CHIP mutations. IL- 1RAP is a transmembrane protein that potentiates multiple inflammatory signaling pathways, including IL-1, IL-33, IL-36G, and stem cell factor (27, 28), and it has a unique feature of being expressed at higher levels in stem and progenitor cells from myeloid leukemia patients compared to normal HSPC (50–53). These properties of IL-1RAP led to several studies investigating the targetability of IL-1RAP as a treatment strategy for myeloid leukemia (25, 52, 54, 55) and may underlie its modification effects of CHIP-associated CVD and, potentially, other disease risks.

These observations agree with the aforementioned human genetic observations using a common missense variant in *IL6R* (7). Furthermore, *Dnmt3a*-inactivated lineage-negative bone marrow cells versus wild-type cells transplanted into mice had greater IL-6 concentrations (56), and humans with *DNMT3A* mutations had greater expression of NLRP3-related cytokines among peripheral blood mononuclear cells (18). While the results above and a prior murine study support the role of AIM2 in *JAK2* CHIP, IL-1β inhibiton was shown to also influence indices related to plaque stability in *Jak2*^VF^ transgenic mice (15). Given the significant impact of IL- 1RAP predicted expression across all CHIP-associated CAD risks, whether IL-1RAP represents a more effective therapeutic target than individual inflammasomes or their downstream effectors warrants further study.

Finally, our approach of using genetically predicted expression as therapeutic instruments in humans can potentially advance precision medicine for CVD and beyond.

Precision medicine aims to identify and implement therapies that are maximally efficacious based on key features (57). We leverage prior insights showing the value of human genetics for therapeutic development prioritization (19). Prior studies have similarly used genotype-imputed transcriptomics to nominate therapeutic targets (58–60). Given the overall relatively low heritability of inflammatory gene expression, we use both summary- and individual-level training data to impute gene expression perturbations from human genetics. We now compare effects by strata to identify subgroups that may maximally clinically benefit from inflammation modulation. Our subsequent murine validation lends overall support to this framework.

Our study has important limitations. First, the predicted expression scores for inflammatory genes are genetic proxies for expression levels from birth, which is well before the acquisition of age-related CHIP mutations. Thus, our analyses do not capture the modification effects after CHIP is manifest, which would more closely mimic clinical trials. However, our approach was corroborated by modeling in murine macrophages introducing an inflammatory stimulus after a CHIP mutation is introduced. Second, CHIP mutations remain uncommon in the unselected population, so power is limited for interaction analyses. Third, our framework is similarly dependent on suitable heritabilities of the gene expression instruments, and we are thus underpowered to detect associations for instruments with low heritabilities. Since we used individual-level validation data, we were able to exclude instruments with very low heritabilities to optimize multiple hypothesis testing. Fourth, the majority of participants in our study population, as well as the eQTLGen Consortium, which we used for generating the predicted expression score, were of European ancestry (20, 61); therefore, our findings may not be generalizable to other ancestries. Finally, our computational approaches using human genetics discovered potential modifications of *ASXL1*-associated CVD risk, with is supported by our experiments using *Asxl1* mutant BMDMs. We set out to model *Asxl1* clonal hematopoiesis in vivo and monitor atherosclerosis. Yet, in line with other studies (43), we found that introducing *Asxl1* mutations via bone marrow transplantation in mice did not confer a clonal advantage or lead to the development of atherosclerosis within a short timeframe. Further research is required to establish a more suitable model prior to drawing conclusions.

In conclusion, validating the approach used, our study replicated murine findings in humans that *JAK2* CHIP mutation enhances CVD risk and the unexpected finding that genetically reduced *Aim2* expression specifically reduces this risk. Examinations across the modification of other CHIP mutation-associated CVD risk by the predicted expression levels of other inflammatory genes yield several novel findings, including modification by *AIM2* expression in *ASXL1*-associated CVD risk, which we corroborated in CRISPR-induced *Asxl1* murine macrophages. Our results may contribute to developing CHIP-type specific CVD therapies and advance precision medicine goals.

## Methods

### Study Population

In the current analysis, we included the first 424,651 unrelated participants enrolled in the UK Biobank study who underwent exome sequencing of blood DNA and were free of hematologic cancer and CVD at baseline (62, 63). Between 2006 and 2010, approximately 500,000 residents of the UK aged 40-69 years were recruited at one of 22 assessment centers across the UK and had samples, including blood-derived DNA, collected at baseline as well as baseline clinical characteristics, biomarkers, and subsequently incident clinical events through medical history and linkage to data on hospital admissions and mortality.

Details regarding this cohort have been described elsewhere in detail (61). Relatedness was defined as one individual in each pair within the third degree of relatedness determined based on kinship coefficients centrally calculated by UK Biobank (61).

### Whole exome sequencing and CHIP detection

Exomes of approximately 450,000 UK Biobank participants were sequenced from blood-derived DNA at the Regeneron Genetics Center, as reported previously (63). Briefly, exomes were captured by IDT’s xGen probe library and sequenced on the Illumina Novaseq platform. Sample-specific FASTQ. files were aligned to the GRCh38 reference. The resultant binary alignment file (BAM) containing the genomic information was evaluated for duplicate reads using Picard3 MarkDuplicates tool and then converted by samtools to CRAM files that, after going through quality controls, were submitted to the UK Biobank data repository for distribution. CHIP detection was conducted through using GATK Mutect2 software (https://software.broadinstitute.org/gatk) as previously performed (7, 64, 65). Participants were annotated as having putative CHIP if the output contained at least one of a pre-specified list of putative CHIP variants in 74 genes anticipated to cause myeloid malignancy at a VAF>2% (Supplemental Table 7) (3, 6, 66). Common sequencing artifacts and germline variants were excluded, as described elsewhere (7).

### RNA sequencing data

RNA sequencing (RNA-seq) data was obtained from two TransOmics in Precision Medicine (TOPMed) cohorts: Multi-Ethnic Study of Atherosclerosis (MESA) and Framingham Heart Study (FHS).

MESA is a multi-ancestry prospective cohort of 6,814 self-identified White, Black, Hispanic, or Asian men and women free of clinical cardiovascular disease at recruitment in 2000-2002(67). Included in this study are 889 individuals who had RNA-seq data in peripheral blood mononuclear cells (PBMCs) measured at baseline. A total of 889 participants were randomly selected from the MESA cohort for RNA sequencing for PBMC following standard protocol. The technical details for the sample acquisition and RNA sequencing can be found at Liu et al. (68).

FHS is a multi-generational cohort initiated in 1948(69). The Framingham Offspring cohort (Gen 2) was recruited in 1971 (N=5,124), and the Generation 3 (Gen 3) cohort was recruited in 2002-2005 (N=4,095) (70, 71). The participants were predominantly self-identified White. Included in this study are 2,622 individuals from the Offspring and Gen 3 cohorts who had their peripheral whole blood samples collected and blood RNA sequenced at exams 9 and 2, respectively. The technical details for the blood draw and RNA sequencing can be found at Liu et al. (72).

### Gene selection and predicted expression score generation

We examined pairs of common CHIP mutations that are associated with CVD risk(6), including *DNMT3A*, *TET2*, *ASXL1*, and *JAK2*, and genetically-predicted expression levels of inflammatory genes that are biologically closely related to the NLRP3 or AIM2 inflammasomes; these genes were selected based on established biological pathways (73, 74) and protein-protein interactions (75). Specifically, both regulated by IFN-γ (73, 76), AIM2, and NLRP3 inflammasome activation lead to cleavage of IL- 1β and IL-18 to produce their mature forms(77, 78). IL-1β and IL-18 in their active forms then exert diverse biological functions related to inflammation(79), including inducing the production of IL-6, a strong independent predictor of cardiovascular outcomes (80, 81). We, therefore, include genes encoding these key proteins, namely *IFNG*, *AIM2*, *NLRP3*, *IL1B*, *IL18*, and *IL6R*. Based on the protein-protein interaction networks provided by STRING (https://string-db.org/), we further extended our study to genes that encode proteins with the top 10 highest interaction scores with each of the key proteins (since AIM2 and NLRP3 highly interact, we only kept one of them, NLRP3, as a key protein for selecting genes in the extended list). This resulted in a total of 29 additional genes, namely *CARD8, CASP1, CASP5, DHX33, IFNGR1, IFNGR2, IL10, IL18BP, IL18R1, IL18RAP, IL1R1, IL1R2, IL1RAP, IL6, IL6ST, IRF1, JAK1, JAK2, JAK3, NEK7, NLRC4, SOCS, STAT1, STAT3, STAT4, STAT5A, STAT6, TNF,* and *TYK2*.

For all selected genes, we used genotyping array data from the UK Biobank participants to generate predicted expression scores. The details on quality control and imputation of genotypic data in UK Biobank have been described elsewhere in detail (61). Briefly, genotypic data were obtained using either UK BiLEVE Axiom arrays (Affymetrix Research Service Laboratory) or UK Biobank Axiom and then imputed to either the Haplotype Reference Consortium (HRC) or the merged UK10K+1000 Genomes as reference panel. Principal component analysis (PCA) was performed using fastPCA(82) based on a pruned set of 147,604 single nucleotide variations (SNVs) among unrelated individuals(83).

We calculated the predicted expression score as weighted sums of expression-increase allele counts among selected single nucleotide polymorphisms (SNPs), weighted by their raw or posterior effect sizes on the expression levels of the corresponding genes (beta coefficient) (22, 84). Raw beta coefficient estimates based on summary statistics of the whole blood (85% of the Consortium) and peripheral blood mononuclear cells (PBMC; 15% of the Consortium) cis-eQTL results from the eQTLGen Consortium (N: 31,684; https://www.eqtlgen.org/) (20), with *cis* being defined as within +/-500,000 bp around the transcriptional start site (TSS) of the encoding gene of the target protein. The majority of participants included in the eQTLGen Consortium are of European descent, which is similar to our study population (20). We used two methods to calculate the scores among EA and non-EA participants separately. (1) The pruning + thresholding (P+T) approach, where we used the raw effect size as weights for SNPs and conducted SNPs selection based on the following:

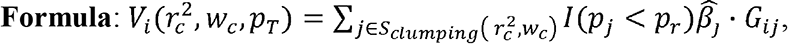

where for individual i, 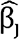 and p are the effect size and *P* of variant j estimated from the summary statistics, respectively, G_ij_ is the genotype dosage for that individual i and variant j, the set of S_C1umping_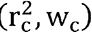 means restricting to variants remained after clumping at the squared correlation threshold of 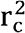 and clumping window size of w, and I (p_j_ <p_r_) is a binary indicator function with 1 indicating *P* of variant j less than the specific *P* cutoff p_r_ and 0 the other way(21).

For each gene, we created 30 candidates P+T-based predicted expression scores based on three *r*^2^ levels (0.1, 0.01, and 0.001), five *P*Lvalue thresholds (5L×L10^−8^, 1L×L10^−5^, 0.001, 0.01, and 0.1), and two clumping window size (within 250kb and 5mb to both ends of the index SNP). (2) The PRS-CS approach, which uses a continuous shrinkage Bayesian framework to calculate the posterior mean of effect sizes, used as weights, across all SNPs (22). For each gene, we also created four candidate PRS-CS-based predicted expression scores using four candidate global shrinkage parameters (1 ×L10^−6^, 1L×L10^−4^, 0.01, and 1). For both approaches, we used a set of unrelated individuals from phase 3 1000 Genomes Project as the LD reference panel (85). Since eQTLGen summary statistics were from both whole bold and peripheral blood mononuclear cells (PBMC) samples, we used genotypes and transcriptome concentrations from both FHS (whole blood) and MESA (PBMC) for score tunning (68). For each gene, we selected the optimal method and parameters for generating the score based on the largest R^2^ of the measured transcriptome levels in either FHS or MESA, since the eQTL source data was from either whole blood or PBMC. The best-predicted expression scores were all standardized to zero-mean and unit-variance and were approximately normally distributed in the population. In the current study, we kept genes whose final-selected best-performed predicted expression scores had R^2^ >1% among EA participants, resulting in suitable scores for 26 genes (**Figure 2** and **Supplemental Table 2**).

### Study outcomes

The primary outcome, CVD event, was a composite of composite of myocardial infarction, coronary artery revascularization, stroke, or death as before(7). We also secondarily used CAD for sensitivity analysis, which was defined as myocardial infarction, percutaneous transluminal coronary angioplasty or coronary artery bypass grafting, chronic ischemic heart disease, and angina. Both disease outcomes were defined by a combination of inpatient hospital billing International Classification of Diseases (ICD) codes and UK death registries listed in **Supplemental Table 8** (7). The exploratory outcomes included 31 hematopoietic cell count indices and indices and five cardiometabolic biomarkers (C-reactive protein [CRP], total cholesterol, high-density lipoprotein [HDL] cholesterol, low-density lipoprotein [LDL] cholesterol, and triglycerides). These conventionally measured biomarkers were analyzed as quantitative traits and were log2-transformed (with adding one across all measurements to avoid zero values for CRP), standardized to zero-mean and unit-variance, and normalized in the population. Blood samples of UK Biobank participants were collected into 4- mL EDTA vacutainers by vacuum draw, stored at 4 degrees centigrade, and then transported to the UK Biocentre in temperature-controlled shipping boxes (86). Full blood counts were measured among all participants using clinical hematology analyzers at the centralized processing laboratory (Stockport, UK). Serum CRP level was measured by immunoturbidimetric high-sensitivity analysis on a Beckman Coulter AU5800. Lipid measurements were performed on the Beckman Coulter AU5800 platform and run using an immune-turbidimetric approach.

### Asxl1-chimeric mice

Bone marrow from CD45.2^+^ Cas9 transgenic mice (The Jackson Laboratory, 026179) was harvested and enriched for c-Kit^+^ cells using magnetic beads (Miltenyi Biotec, Cat # 130-091-224). LT-HSCs (Lin^-^c-Kit^+^Sca1^+^CD48^-^CD150^+^) (87) were then harvested by flow cytometry sorting. LT-HSCs were then spinfected with 6ug/mL Polybreen (MilliporeSigma, TR-1003-G) and Lentiviruses containing non-targeting guides (Nmt4) or guides targeted to Asxl1 in exon 12 (*Asxl1*-G623*). LT-HSCs were washed and then incubated for three days. LT-HSCs were then mixed with 1 million supporting cells from CD45.1^+^ WT mice and transplanted into irradiated *Ldlr^-/-^* recipient mice.

### Asxl1-CRISPR validation

CRISPR guides designed to exon 12 of *Asxl1* were designed by CHOPCHOP (88) and screened in skin-derived fibroblasts from Cas9 transgenic mice. Guide sequence AGTGGTAACCTCTCGCCCCTCGG was evaluated by sanger sequencing of PCR amplification of flanking regions using forward GCAGCATAAAATGGCTCTTGAT and reverse GCTGAGTCTTCTCTTCTGGCTC primers.

### Inflammasome activation studies

Five weeks after transplantation, bone marrow was harvested and cultured in L-Cell Media for five days to generate bone marrow-derived macrophages (BMDMs). 20,000 BMDMs/well were seeded into 96 well plates and allowed to recover overnight. BMDMs were then primed with 20ng/mL LPS (Cell Signaling, 14011) for three hours and then stimulated with the indicated concentrations of ATP (Sigma) for 1 hour. For AIM2 inflammasome activation BMDMs were primed for 1 hour with 20ng/mL LPS (Cell Signaling, 14011) then incubated with lipofectamine 2000 (Thermofisher, Cat# 11668019) and poly(deoxyadenylic-deoxythymidylic) acid sodium salt (pdAdT) (Invivogen, tlrl-patn) for 6 hours. Following incubations, supernatants were collected, spun down at 3000g for 10 minutes, then assessed for IL-1β protein by ELISA (R&D Systems, DY401) and LDH activity (Thermofisher, Cat# C20301).

### BMDM cultures

For Protein secretion assays, bone marrow was harvested as indicated above, and after five days of differentiation in L cell media, BMDMs were seeded at 20,000/well in 96 well plates and allowed to recover overnight. Cells were then treated with vehicle (PBS) or LPS 20ng/mL final concertation for 6 hours. Media was collected frozen, and ELISA was conducted to determine concentrations of IL-6 (R&D Systems, DY406), TNFα (R&D Systems, DY410), or IL-10 (R&D Systems, DY417).

For mRNA analysis, BMDM were differentiated for five days, then seeded into 12 well plates and allowed to recover overnight. Cells were then treated with vehicle (PBS) or LPS 20ng/mL final concentration for 6 hours. BMDMs were then rinsed 3x with PBS and suspended in Trizol Reagent (ThermoFisher, 15596026), and RNA was isolated using an RNeasy micro kit (Qiagen, 74004) with DNase digestion. cDNA was then synthesized (ThermoFisher, 4368814), and qPCR analysis was conducted and normalized to β-Actin expression.

pγH2AX western blot analysis was conducted on BMDMs differentiated for five days and plated into six-well dishes and allowed to recover overnight. BDMDs were treated with the indicated stimulus, including LPS 20ng/mL for 6 hours. Cells were then washed 3x with PBS, and protein was isolated in RIPA buffer (Boston Bioproducts, BP-115) with protease and phosphatase inhibitors (ThermoFisher, 78439). Protein was quantified with BCA analysis and subjected to western blot using antibodies to pγH2AX (Cell Signaling, 9718) and β-Actin (Cell Signaling, 12262).

### Atherosclerosis studies

Bone marrow transplantations were conducted as described above into lethally irradiated *Ldlr^-/-^* mice. After four weeks of recovery, mice were subjected to western- type diet (WTD) feeding for 12 weeks. Blood cell counts were quantified from cheek bleeding using a VetScan HM5 Hematology system (Abaxis). For Asxl1 burden analysis, red blood cells were lysed using RBC lysis buffer (Biolegend, 420301), washed in PBS with 1% BSA and 2mM EDTA, stained with the indicated antibodies (CD3, CD115, Ly6G, CD45.1, and CD45.2), and then analyzed using a BD Fortessa. After 12 weeks of WTD feeding, mice were euthanized, perfused with PBS, and aortic roots were fixed in 4% paraformaldehyde for 48 hours. Aortic roots were embedded in paraffin and sectioned 6μm thick. Hemotoxin and Eosin staining was conducted on six slides 60μm apart and imaged on a Nikon Labphoto 2 and Image Pro Plus software (Media Cybernetics version 7.0.0.591). Blinded researchers quantified lesion area and necrotic core area in FIJI software (89) with an average of the six slides reported.

### Statistics

We evaluated the association between CHIP mutations and incident CVD, as well as the modification effects by predicted expression levels of inflammatory genes measured as predicted expression scores. Using Cox proportional hazard models, we first estimated the hazard ratios (HRs) and associated 95% confidence intervals (CI) of (1) the presence of CHIP mutations and (2) the presence of large clones, defined as a variant allele fraction (VAF) >10%, of CHIP mutations for incident CVD events. Then we conducted stratified analyses evaluating the associations between the predicted expression scores of selected inflammatory genes on the incidence of the primary outcome (i.e., CVD) with or without the presence of CHIP variables. For predicted expression scores that were associated with incident CVD risk (defined as *P*<0.05) only in the presence of CHIP variables(s), we carried forward to evaluate the interactions between those scores and the corresponding CHIP variables on the primary outcome. We considered time at risk to start at the enrollment of the study and continue until the event of interest, death, loss to follow-up, or the end of follow-up. Models were adjusted for age at the time of enrollment, sex, self-reported white British race/ethnicity, BMI, diagnoses of type 2 diabetes mellitus at the time of enrollment, ever-smoker status, and the first ten principal components of genetic ancestry. Since only less than 2% of the study population has missingness for any of the adjusted covariates, we removed those individuals from our regression models.

For significant interactions (FDR <0.05) discovered in the above analysis, we evaluated their associations across 31 hematological and five cardiometabolic traits using the same Cox proportional hazard models with adjusting for the same sets of covariates. All hematological and lipid traits were log2-transformed, standardized to zero-mean and unit-variance, and were approximately normally distributed in the population. Analyses used R version 4.0.0 software (The R Foundation, Vienna, Austra), two-tailed *P*-values, as well a statistical significance level of 0.05 for other analyses.

### Study approval

The secondary use of data for the present analysis was approved by the Massachusetts General Hospital Institutional Review Board (protocol 2021P002228) and facilitated through UK Biobank Application 7089.

## Author contributions

Z.Y., T.P.F., A.R.T., and P.N. conceptualized the study. Z.Y., T.P.F., Y.R., A.R.T., and P.N. developed the methodology. Z.Y., T.P.F., and Y.R. performed human statistical analysis and mice experiments. C.V., T.N., M.M.U., T.M., A.N., J.B.H., C.J.G., and G.K.G. conducting CHIP calling. S.M.Z. curated the phenotypes and covariates. Y.W., G.M.P., N.H., D.L., R.S.V., F.A., K.A., K.D.T., S.S.R., and J.I.R. generated and managed the RNA-sequencing data used for eQTL score validation. P.L., S.J., B.L.E., A.G.B., A.R.T., and P.N. supervised the research. A.L.T. and P.N. acquired the funding for the research. Z.Y. and T.F. wrote the original draft manuscript. Y.R., C.V., T.N., M.M.U., T.M., A.N., J.B.H., S.M.Z., C.J.G., G.K.G., Y.W., G.M.P., N.H., D.L., R.S.V., F.A., K.A., K.D.T., S.S.R., J.I.R., P.L., S.J., B.L.E., A.G.B., A.R.T., and P.N. reviewed and edited the manuscript.

## Data Availability

UK Biobank individual-level data are available by request via the application at https://www.ukbiobank.ac.uk. TOPMed individual-level DNA and RNA sequencing data used in this analysis are available through restricted access via the dbGaP.

## Acknowledgments

Whole genome sequencing (WGS) for the Trans-Omics in Precision Medicine (TOPMed) program was supported by the National Heart, Lung and Blood Institute (NHLBI). WGS for “NHLBI TOPMed: Multi-Ethnic Study of Atherosclerosis (MESA)” (phs001416.v1.p1) was performed at the Broad Institute of MIT and Harvard (3U54HG003067-13S1). Centralized read mapping and genotype calling, along with variant quality metrics and filtering were provided by the TOPMed Informatics Research Center (3R01HL-117626-02S1). Phenotype harmonization, data management, sample-identity QC, and general study coordination, were provided by the TOPMed Data Coordinating Center (3R01HL-120393-02S1), and TOPMed MESA Multi-Omics (HHSN2682015000031/HSN26800004). The MESA projects are conducted and supported by the National Heart, Lung, and Blood Institute (NHLBI) in collaboration with MESA investigators. Support for the Multi-Ethnic Study of Atherosclerosis (MESA) projects are conducted and supported by the National Heart, Lung, and Blood Institute (NHLBI) in collaboration with MESA investigators. Support for MESA is provided by contracts 75N92020D00001, HHSN268201500003I, N01-HC-95159, 75N92020D00005, N01-HC-95160, 75N92020D00002, N01-HC-95161, 75N92020D00003, N01-HC-95162, 75N92020D00006, N01-HC-95163, 75N92020D00004, N01-HC-95164, 75N92020D00007, N01-HC-95165, N01- HC-95166, N01-HC-95167, N01-HC-95168, N01-HC-95169, UL1-TR-000040, UL1-TR- 001079, UL1-TR-001420, UL1TR001881, DK063491, and R01HL105756. The authors thank the other investigators, the staff, and the participants of the MESA study for their valuable contributions. A fill list of participating MESA investigators and institutes can be found at http://www.mesa-nhlbi.org. This study was also supported in part by the National Institutes of Health, National Heart, Lung, Long and Blood Institute (NHLBI) contract 1R01HL151855 and the National Institute of Diabetes and Digestive and Kidney Diseases contract UM1DK078616. The Framingham Heart Study (FHS) acknowledges the support of contracts NO1-HC-25195, HHSN268201500001I and 75N92019D00031 from the National Heart, Lung and Blood Institute and grant supplement R01 HL092577-06S1 for this research. We also acknowledge the dedication of the FHS study participants without whom this research would not be possible. Dr. Vasan is supported in part by the Evans Medical Foundation and the Jay and Louis Coffman Endowment from the Department of Medicine, Boston University School of Medicine.

