## Supplemental Material for "Genetic modification of inflammation and clonal hematopoiesis-associated cardiovascular risk"

**Supplemental Table 1. Associations between CHIP mutation and incidence of CAD event**. CAD event outcome encompasses individuals with myocardial infarction, percutaneous transluminal coronary angioplasty or coronary artery bypass grafting, chronic ischemic heart disease, and angina. Models were adjusted for age at the time of enrollment, sex, white British ancestry, body mass index, diagnoses of type 2 diabetes mellitus at the time of enrollment, ever-smoker status, and the first ten principal components of genetic ancestry. Participants with prevalent hematological cancers or CAD were removed from the analyses.

| | **Presence of CHIP** | | **Presence of large CHIP** [a] | |
|---|---|---|---|---|
| | **HR (95% CI)** | **P-value** | **HR (95% CI)** | **P-value** |
| **CHIP** | 1.05 (1.00, 1.10) | 0.05 | 1.09 (1.02, 1.17) | 0.008 |
| ***DNMT3A*** | 0.97 (0.91, 1.04) | 0.37 | 1.01 (0.91, 1.11) | 0.91 |
| ***TET2*** | 1.02 (0.92, 1.13) | 0.67 | 1.18 (1.03, 1.35) | 0.02 |
| ***ASXL1*** | 1.21 (1.07, 1.38) | 0.003 | 1.16 (0.97, 1.39) | 0.10 |
| ***JAK2*** | 1.52 (1.04, 2.21) | 0.03 | 1.55 (1.06, 2.26) | 0.02 |
| ***PPM1D*** | 1.18 (0.93, 1.49) | 0.18 | 1.14 (0.78, 1.67) | 0.49 |
| ***TP53*** | 1.20 (0.92, 1.56) | 0.19 | 1.51 (1.02, 2.24) | 0.04 |
| ***SRSF2*** | 1.46 (1.09, 1.95) | 0.01 | 1.47 (1.02, 2.10) | 0.04 |
| ***SF3B1*** | 1.21 (0.84, 1.76) | 0.30 | 1.21 (0.79, 1.86) | 0.38 |

[a] Large CHIP is defined as a variant allele fraction >10%.

CHIP, clonal hematopoiesis of indeterminate potential; CAD, coronary artery disease; HR, hazard ratio

**Supplemental Table 2. Adjusted proportion of the variance for gene expression level explained by the best-performed predicted expression scores generated using P+T or PRS-CS approaches.**

| | EUR | | Non-EUR | |
|---|---|---|---|---|
| **Gene** | **Method** | **Variance** | **Method** | **Variance** |
| *IL18RAP* | PT | 34.70% | PT | 3.34% |
| *IL1R1* | PT | 14.87% | PT | 3.78% |
| *TNF* | PT | 8.97% | PT | 1.96% |
| *IL10* | PT | 8.75% | PT | 4.80% |
| *IL1R2* | PT | 7.48% | PT | 3.30% |
| *AIM2* | PT | 7.33% | PRS-CS | 10.96% |
| *IL18R1* | PRS-CS | 6.36% | PRS-CS | 1.29% |
| *CASP5* | PT | 6.22% | PT | 1.63% |
| *TYK2* | PT | 4.50% | PRS-CS | 0.99% |
| *IL18* | PRS-CS | 4.34% | PRS-CS | 3.44% |
| *CASP1* | PT | 4.27% | PT | 2.14% |
| *NLRP3* | PT | 4.16% | PRS-CS | 0.95% |
| *JAK3* | PRS-CS | 3.57% | PT | 2.39% |
| *CARD8* | PRS-CS | 3.52% | PT | 10.82% |
| *NLRC4* | PT | 3.05% | PT | 1.59% |
| *JAK2* | PT | 3.03% | PRS-CS | 2.35% |
| *IFNGR2* | PT | 2.76% | PRS-CS | 5.13% |
| *IFNGR1* | PRS-CS | 2.04% | PRS-CS | 2.23% |
| *IL18BP* | PT | 1.84% | PRS-CS | 0.50% |
| *IL6* | PT | 1.82% | PT | 1.50% |
| *IL1RAP* | PRS-CS | 1.69% | PT | 1.96% |
| *IL6ST* | PT | 1.53% | PRS-CS | 0.55% |
| *STAT4* | PRS-CS | 1.32% | PT | 1.00% |
| *STAT6* | PRS-CS | 1.32% | PRS-CS | 1.55% |
| *NEK7* | PT | 1.15% | PT | 0.58% |
| *IL1B* | PT | 1.05% | PRS-CS | 0.91% |

PRS-CS: polygenic risk score-continuous shrinkage; P+T: pruning and thresholding.

**Supplemental Table 3. Associations between predicted expression scores of inflammatory genes on incident CVD events stratified by CHIP genes.** CVD event outcome is defined as a composite of myocardial infarction, coronary artery disease or revascularization, stroke, or death. Models were adjusted for age at the time of enrollment, sex, white British ancestry, BMI, diagnoses of type 2 diabetes mellitus at the time of enrollment, ever-smoker status, and the first ten principal components of genetic ancestry.

| **CHIP variables** | **CHIP** | | ***DNMT3A*** | | ***TET2*** | | ***ASXL1*** | | ***JAK2*** | |
|---|---|---|---|---|---|---|---|---|---|---|
| | **Not present (N= 385,678)** | **Present (N= 25,344)** | **Not present (N= 396,965)** | **Present (N= 14,057)** | **Not present (N= 405,978)** | **Present (N= 5,044)** | **Not present (N= 408,719)** | **Present (N= 2,303)** | **Not present (N= 410,776)** | **Present (N= 246)** |
| ***AIM2*** | 0.99 (0.98, 1.00)[a] | 1.00 (0.97, 1.03) | 0.99 (0.98, 1.00) | 1.00 (0.95, 1.05) | 0.99 (0.98, 1.00) | 1.00 (0.93, 1.08) | 0.99 (0.98, 1.00) | 0.95 (0.87, 1.03) | 0.99 (0.98, 1.00) | 1.85 (1.12, 3.07) |
| ***CARD8*** | 1.01 (1.00, 1.02) | 0.99 (0.96, 1.02) | 1.01 (1.00, 1.02) | 0.98 (0.94, 1.02) | 1.01 (1.00, 1.02) | 1.00 (0.93, 1.07) | 1.01 (1.00, 1.02) | 1.03 (0.94, 1.13) | 1.01 (1.00, 1.02) | 1.06 (0.84, 1.34) |
| ***CASP1*** | 1.00 (0.99, 1.01) | 1.01 (0.98, 1.04) | 1.00 (0.99, 1.01) | 0.99 (0.95, 1.04) | 1.00 (0.99, 1.01) | 1.01 (0.94, 1.08) | 1.00 (0.99, 1.01) | 1.03 (0.94, 1.13) | 1.00 (0.99, 1.01) | 0.94 (0.72, 1.23) |
| ***CASP5*** | 1.00 (0.99, 1.01) | 0.99 (0.96, 1.02) | 1.00 (0.99, 1.01) | 0.98 (0.94, 1.03) | 1.00 (0.99, 1.01) | 1.00 (0.93, 1.07) | 1.00 (0.99, 1.01) | 1.03 (0.94, 1.13) | 1.00 (0.99, 1.01) | 1.08 (0.86, 1.36) |
| ***IFNGR1*** | 1.00 (0.99, 1.01) | 0.98 (0.95, 1.01) | 1.00 (0.99, 1.01) | 0.99 (0.95, 1.04) | 1.00 (0.99, 1.01) | 0.99 (0.92, 1.06) | 1.00 (0.99, 1.01) | 1.04 (0.95, 1.14) | 1.00 (0.99, 1.01) | 1.29 (1.01, 1.66) |
| ***IFNGR2*** | 1.00 (0.99, 1.01) | 1.02 (0.98, 1.05) | 1.00 (0.99, 1.01) | 1.01 (0.96, 1.06) | 1.00 (0.99, 1.01) | 1.00 (0.94, 1.08) | 1.00 (0.99, 1.01) | 1.00 (0.91, 1.10) | 1.00 (0.99, 1.01) | 0.96 (0.74, 1.25) |
| ***IL10*** | 1.00 (0.99, 1.01) | 0.99 (0.96, 1.02) | 1.00 (0.99, 1.01) | 0.99 (0.95, 1.04) | 1.00 (0.99, 1.01) | 1.03 (0.96, 1.11) | 1.00 (0.99, 1.01) | 0.91 (0.83, 0.99) | 1.00 (0.99, 1.01) | 1.01 (0.81, 1.26) |
| ***IL18*** | 1.01 (1.00, 1.02) | 0.99 (0.96, 1.02) | 1.01 (1.00, 1.02) | 0.97 (0.93, 1.02) | 1.00 (1.00, 1.01) | 0.96 (0.90, 1.03) | 1.00 (0.99, 1.01) | 1.03 (0.95, 1.13) | 1.00 (0.99, 1.01) | 0.95 (0.75, 1.20) |
| ***IL18BP*** | 0.99 (0.98, 1.00) | 1.01 (0.98, 1.04) | 0.99 (0.99, 1.00) | 1.01 (0.97, 1.06) | 0.99 (0.99, 1.00) | 1.03 (0.96, 1.12) | 1.00 (0.99, 1.01) | 0.98 (0.89, 1.07) | 1.00 (0.99, 1.00) | 1.10 (0.80, 1.53) |
| ***IL18R1*** | 1.00 (0.99, 1.01) | 0.98 (0.95, 1.02) | 1.00 (0.99, 1.01) | 1.00 (0.95, 1.04) | 1.00 (0.99, 1.01) | 0.98 (0.91, 1.05) | 1.00 (0.99, 1.01) | 0.99 (0.91, 1.08) | 1.00 (0.99, 1.01) | 1.14 (0.90, 1.44) |
| ***IL18RAP*** | 1.00 (0.99, 1.01) | 0.99 (0.96, 1.02) | 1.00 (0.99, 1.01) | 1.01 (0.97, 1.06) | 1.00 (0.99, 1.01) | 1.00 (0.93, 1.07) | 1.00 (0.99, 1.01) | 0.90 (0.83, 0.98) | 1.00 (0.99, 1.01) | 1.11 (0.86, 1.42) |
| ***IL1B*** | 1.01 (1.00, 1.02) | 1.03 (1.00, 1.06) | 1.01 (1.00, 1.02) | 1.05 (1.01, 1.09) | 1.01 (1.00, 1.02) | 1.03 (0.97, 1.09) | 1.01 (1.00, 1.02) | 1.00 (0.91, 1.10) | 1.01 (1.00, 1.02) | 0.95 (0.69, 1.31) |
| ***IL1R1*** | 0.99 (0.98, 1.00) | 1.00 (0.97, 1.03) | 0.99 (0.98, 1.00) | 0.99 (0.95, 1.03) | 0.99 (0.98, 1.00) | 0.95 (0.88, 1.02) | 0.99 (0.98, 1.00) | 1.02 (0.93, 1.11) | 0.99 (0.98, 1.00) | 1.05 (0.82, 1.35) |
| ***IL1R2*** | 1.00 (0.99, 1.01) | 1.01 (0.98, 1.04) | 1.00 (0.99, 1.01) | 1.00 (0.95, 1.05) | 1.00 (0.99, 1.01) | 0.98 (0.91, 1.05) | 1.00 (0.99, 1.01) | 0.94 (0.86, 1.03) | 1.00 (0.99, 1.01) | 1.19 (0.88, 1.60) |
| ***IL1RAP*** | 1.00 (0.99, 1.01) | 1.04 (1.01, 1.07) | 1.00 (0.99, 1.01) | 1.06 (1.02, 1.11) | 1.00 (0.99, 1.01) | 1.04 (0.97, 1.11) | 1.00 (0.99, 1.01) | 0.97 (0.89, 1.07) | 1.00 (0.99, 1.01) | 1.38 (1.13, 1.69) |
| ***IL6*** | 1.00 (0.99, 1.01) | 0.99 (0.95, 1.02) | 1.00 (0.99, 1.01) | 1.02 (0.98, 1.07) | 1.00 (0.99, 1.01) | 0.93 (0.87, 1.00) | 1.00 (0.99, 1.01) | 0.99 (0.90, 1.08) | 1.00 (0.99, 1.01) | 0.92 (0.73, 1.16) |
| ***IL6ST*** | 1.00 (0.99, 1.01) | 1.02 (0.98, 1.05) | 1.00 (0.99, 1.01) | 1.01 (0.97, 1.06) | 1.00 (0.99, 1.01) | 1.07 (1.00, 1.15) | 1.00 (0.99, 1.01) | 1.00 (0.92, 1.10) | 1.00 (0.99, 1.01) | 1.10 (0.88, 1.38) |
| ***JAK2*** | 1.01 (1.00, 1.02) | 1.01 (0.98, 1.04) | 1.01 (1.00, 1.02) | 1.01 (0.96, 1.05) | 1.01 (1.00, 1.02) | 1.04 (0.96, 1.11) | 1.01 (1.00, 1.02) | 0.99 (0.91, 1.08) | 1.01 (1.00, 1.02) | 1.00 (0.81, 1.23) |
| ***JAK3*** | 1.00 (0.99, 1.01) | 1.00 (0.97, 1.03) | 1.00 (0.99, 1.01) | 1.03 (0.98, 1.08) | 1.01 (1.00, 1.01) | 0.96 (0.89, 1.03) | 1.00 (0.99, 1.01) | 0.99 (0.91, 1.08) | 1.00 (0.99, 1.01) | 0.96 (0.77, 1.20) |
| ***NEK7*** | 0.99 (0.98, 1.00) | 1.00 (0.97, 1.03) | 0.99 (0.98, 1.00) | 1.01 (0.96, 1.06) | 0.99 (0.98, 1.00) | 0.98 (0.91, 1.06) | 0.99 (0.98, 1.00) | 0.96 (0.88, 1.06) | 0.99 (0.98, 1.00) | 1.02 (0.82, 1.26) |
| ***NLRC4*** | 0.99 (0.98, 1.00) | 0.97 (0.94, 1.01) | 0.99 (0.98, 1.00) | 0.96 (0.92, 1.01) | 0.99 (0.98, 1.00) | 1.01 (0.94, 1.09) | 0.99 (0.98, 1.00) | 0.97 (0.89, 1.05) | 0.99 (0.98, 1.00) | 0.94 (0.73, 1.22) |
| ***NLRP3*** | 1.00 (0.99, 1.01) | 1.01 (0.98, 1.04) | 1.00 (0.99, 1.01) | 1.00 (0.95, 1.04) | 1.00 (0.99, 1.01) | 1.01 (0.94, 1.08) | 1.00 (0.99, 1.01) | 1.00 (0.92, 1.10) | 1.00 (0.99, 1.01) | 1.20 (0.93, 1.54) |
| ***STAT4*** | 1.00 (0.99, 1.01) | 0.99 (0.96, 1.03) | 1.00 (0.99, 1.01) | 1.01 (0.96, 1.05) | 1.00 (0.99, 1.01) | 0.99 (0.92, 1.06) | 1.00 (0.99, 1.01) | 0.97 (0.88, 1.06) | 1.00 (0.99, 1.01) | 0.92 (0.72, 1.19) |
| ***STAT6*** | 0.99 (0.98, 1.00) | 0.97 (0.94, 1.01) | 0.99 (0.98, 1.00) | 0.98 (0.94, 1.02) | 0.99 (0.98, 1.00) | 0.98 (0.92, 1.05) | 0.99 (0.98, 1.00) | 0.97 (0.89, 1.06) | 0.99 (0.98, 1.00) | 1.02 (0.81, 1.30) |
| ***TNF*** | 0.98 (0.97, 0.99) | 0.98 (0.95, 1.02) | 0.98 (0.97, 0.99) | 0.98 (0.93, 1.02) | 0.98 (0.97, 0.99) | 0.97 (0.90, 1.04) | 0.98 (0.97, 0.99) | 0.99 (0.91, 1.08) | 0.98 (0.97, 0.99) | 1.08 (0.84, 1.37) |
| ***TYK2*** | 1.00 (0.99, 1.01) | 1.00 (0.96, 1.03) | 1.00 (0.99, 1.01) | 1.01 (0.96, 1.05) | 1.00 (0.99, 1.01) | 1.04 (0.97, 1.11) | 1.00 (0.99, 1.01) | 0.99 (0.90, 1.09) | 1.00 (0.99, 1.01) | 0.98 (0.75, 1.28) |

[a] Effect estimates are represented as hazard ratio (95% confidence interval).

CHIP, clonal hematopoiesis of indeterminate potential; CVD, cardiovascular disease

**Supplemental Table 4. Z-scores of interactions between predicted expression scores of inflammatory genes and CHIP mutations on incident CVD events.** CVD event outcome is defined as a composite of myocardial infarction, coronary artery disease or revascularization, stroke, or death. Models were adjusted for age at the time of enrollment, sex, white British ancestry, BMI, diagnoses of type 2 diabetes mellitus at the time of enrollment, ever-smoker status, and the first ten principal components of genetic ancestry.

| | ***CHIP*** | ***DNMT3A*** | ***ASXL1*** | ***JAK2*** |
|---|---|---|---|---|
| ***AIM2*** | - | - | 2.38 | 2.21 |
| ***IFNGR1*** | - | - | - | 1.39 |
| ***IL10*** | - | - | -1.98 | - |
| ***IL18RAP*** | - | - | -2.36 | - |
| ***IL1B*** | - | 1.66 | - | - |
| ***IL1RAP*** | 2.44 | 2.72 | - | 2.31 |

CHIP, clonal hematopoiesis of indeterminate potential; CVD, cardiovascular disease

**Supplemental Table 5. Z-scores of associations between predicted expression scores of inflammatory genes and hematopoietic traits and cardiometabolic biomarkers in the presence of CHIP mutations.** Traits were analyzed as quantitative traits and log2-transformed, standardized to zero-mean and unit-variance, and normalized in the population. Models were adjusted for age at the time of enrollment, sex, white British ancestry, BMI, diagnoses of type 2 diabetes mellitus at the time of enrollment, ever-smoker status, and the first ten principal components of genetic ancestry.

| | ***IL1RAP*** **(*DNMT3A* carriers)** | ***IL1RAP*** **(CHIP carriers)** | ***AIM2*** **(*ASXL1* carriers)** | ***IL18RAP*** **(*ASXL1* carriers)** | ***IL1RAP*** **(*JAK2* carriers)** | ***AIM2*** **(*JAK2* carriers)** |
|---|---|---|---|---|---|---|
| **White blood cell (leukocyte) count** | 0.22 | 1.14 | -0.37 | -0.29 | 0.64 | 0.31 |
| **Red blood cell (erythrocyte) count** | 0.17 | -0.22 | -0.82 | -0.06 | -1.29 | 0.45 |
| **Hemoglobin concentration** | 1.07 | 0.38 | -1.03 | 0.42 | -0.84 | -1.07 |
| **Hematocrit percentage** | 1.18 | 0.26 | -0.74 | 0.13 | -0.62 | -0.70 |
| **Mean corpuscular volume** | 1.48 | 0.78 | 0.27 | 0.31 | 1.21 | -1.39 |
| **Mean corpuscular hemoglobin** | 1.04 | 0.77 | -0.07 | 0.62 | 0.81 | -1.42 |
| **Mean corpuscular hemoglobin concentration** | -0.44 | 0.20 | -0.62 | 0.74 | -0.56 | -0.85 |
| **Red blood cell (erythrocyte) distribution width** | -0.67 | -0.40 | 0.23 | -0.10 | 0.59 | -0.12 |
| **Platelet count** | 0.15 | 0.12 | 0.95 | -0.52 | 0.90 | 0.40 |
| **Platelet crit** | 0.13 | -0.28 | 1.20 | -0.74 | 1.09 | 0.17 |
| **Mean platelet volume** | 0.27 | -0.66 | 0.56 | 0.13 | 0.16 | -0.23 |
| **Platelet distribution width** | -0.22 | -0.52 | 1.13 | 0.27 | 0.23 | 0.72 |
| **Lymphocyte count** | 0.20 | 0.21 | -0.78 | -0.95 | 0.09 | -0.61 |
| **Monocyte count** | 1.06 | 1.52 | 0.98 | -1.18 | -0.35 | 1.69 |
| **Neutrophil count** | -0.05 | 0.94 | -0.14 | 0.82 | 0.57 | -0.16 |
| **Eosinophil count** | 0.69 | 0.95 | -0.11 | -3.04 | 0.08 | -0.91 |
| **Basophil count** | 0.08 | 0.01 | 0.36 | 0.32 | -1.08 | 0.41 |
| **Nucleated red blood cell count** | 0.20 | 1.79 | -0.50 | -0.51 | 2.15 | 0.53 |

| | | | | | | |
|---|---|---|---|---|---|---|
| **Lymphocyte percentage** | -0.10 | -1.01 | -0.41 | -1.13 | -0.53 | -0.76 |
| **Monocyte percentage** | 0.95 | 0.81 | 1.46 | -0.67 | -0.35 | 2.02 |
| **Neutrophil percentage** | -0.38 | -0.01 | 0.31 | 2.18 | 0.30 | -0.71 |
| **Eosinophil percentage** | -0.15 | -0.22 | 0.07 | -2.93 | -0.26 | -0.90 |
| **Basophil percentage** | -0.04 | -0.58 | 1.14 | 1.53 | -1.74 | 0.07 |
| **Nucleated red blood cell percentage** | -0.35 | 0.15 | -2.63 | -0.16 | -0.67 | 0.63 |
| **Reticulocyte percentage** | -0.38 | -0.14 | 0.20 | 0.15 | 0.85 | -0.26 |
| **Reticulocyte count** | -0.14 | -0.13 | -1.31 | -0.05 | 0.22 | -0.06 |
| **Mean reticulocyte volume** | 0.27 | 0.02 | -0.45 | -0.20 | 0.79 | -0.98 |
| **Mean sphered cell volume** | 1.37 | 0.77 | -0.49 | -0.19 | 1.25 | -0.95 |
| **Immature reticulocyte fraction** | -0.26 | 0.22 | 0.81 | -0.16 | 0.62 | -0.70 |
| **High light scatter reticulocyte percentage** | -0.16 | 0.12 | 0.63 | 0.14 | 0.96 | -0.38 |
| **High light scatter reticulocyte count** | 0.24 | 0.09 | 0.35 | 0.20 | 0.27 | -0.29 |

CHIP, clonal hematopoiesis of indeterminate potential

**Supplemental Table 6. Z-scores of interaction terms formed by pairs of predicted expression scores of inflammatory genes and CHIP showed significant modification effects on CVD risk on five cardiometabolic traits.** Traits were analyzed as quantitative traits and log2-transformed, standardized to zero-mean and unit-variance, and normalized in the population. Models were adjusted for age at the time of enrollment, sex, white British ancestry, BMI, diagnoses of type 2 diabetes mellitus at the time of enrollment, ever-smoker status, and the first ten principal components of genetic ancestry.

| | ***IL1RAP*** **(*DNMT3A* carriers)** | ***IL1RAP*** **(CHIP carriers)** | ***AIM2*** **(*ASXL1* carriers)** | ***IL18RAP*** **(*ASXL1* carriers)** | ***IL1RAP*** **(*JAK2* carriers)** | ***AIM2*** **(*JAK2* carriers)** |
|---|---|---|---|---|---|---|
| **C-reactive protein** | -1.28 | -0.34 | 0.01 | -0.18 | 1.31 | 1.63 |
| **Cholesterol** | -0.35 | -1.13 | -0.28 | 0.01 | -0.57 | -0.57 |
| **HDL cholesterol** | -1.62 | -2.09 | 0.10 | 0.12 | -1.46 | -0.10 |
| **LDL cholesterol** | 0.11 | -0.59 | -0.78 | 0.17 | -0.53 | -0.75 |
| **Triglycerides** | 0.56 | 0.97 | 2.13 | 0.33 | 1.14 | 1.07 |

CHIP, clonal hematopoiesis of indeterminate potential; HDL, high-density lipoprotein; LDL, low-density lipoprotein

**Supplemental Table 7. List of hematopoietic genes and variants queried.**

| Gene name | Reported mutations used for variant calling |
|---|---|
| ***ASXL1*** | Frameshift/nonsense/splice-site p.358-1541 |
| ***ASXL2*** | Frameshift/nonsense/splice-site in p.380-1435 |
| ***BCOR*** | Frameshift/nonsense/splice-site |
| ***BCORL1*** | Frameshift/nonsense/splice-site |
| ***BRAF*** | G464E, G464V, G466E, G466V, G469R, G469E, G469A, G469V, V471F, V472S, L485W, N581S, I582M, I592M, I592V, D594N, D594G, D594V, D594E, F595L, F595S, G596R, L597V, L597S, L597Q, L597R, A598V, T599I, V600M, V600L, V600K, V600R, V600E, V600A, V600G, V600D, K601E, K601N, R603*, W604R, W604G, S605G, S605F, S605N, G606E, G606A, G606V, H608R, H608L, G615R, S616P, S616F, L618S, L618W |
| ***BRCC3*** | Frameshift/nonsense/splice-site |
| ***CBL*** | RING finger missense p.381-421 |
| ***CBLB*** | RING finger missense p.372-412 |
| ***CEBPA*** | Frameshift/nonsense/splice-site |
| ***CREBBP*** | Frameshift/nonsense/splice-site, D1435E, R1446L, R1446H, R1446C, Y1450C, P1476R, Y1482H, H1487Y, W1502C, Y1503D, Y1503H, Y1503F, S1680del |
| ***CSF1R*** | L301F, L301S, Y969C, Y969N, Y969F, Y969H, Y969D |
| ***CSF3R*** | T615A, T618I, truncating c.741-791 |
| ***CTCF*** | Frameshift/nonsense, R377C, R377H, P378A, P378L |
| ***CUX1*** | Frameshift/nonsense |
| ***DNMT3A*** | Frameshift/nonsense/splice-site, F290I, F290C, F290S, G293R, L295P, L295Q, L295V, V296G, V296L, V296M, W297C, W297L, W297R, W297G, G298W, G298R, G298E, W306C, P307S, P307R, P307L, P307T, G308D, I310F, I310L, I310S, I310T, S312F, R326G, R326H, R326L, R326C, R326S, V328A, V328D, V328G, F331V, G332R, G332E, S337A, S337L, S337P, V339A, V339M, V339G, L344Q, L344P, L344R, L347P, L347R, L347Q,S352N,Y365C, R366C, R366P, R366H, R366G, A368D, A368T, A368V, I369N, I369S, V372D, L373Q, A376P, A376T, A376V, R379H, R379L, R379C, R379S, D389N, I407T, I407N, I407S, W409R, A410D, A410T, G413V, F414L, F414I, F414S, F414V, F414C, K468R, E477Q, E477K, V483G, R484Q, R484W, C494Y, C497G, C497R, C497Y, G498E, H506R, G511E, C514Y, Q527H, Q527P, D529N, D529V, D531N, D531Y, Y533C, S535F, S535P, C537G, C537R, C540Y, G543A, G543S, G543C, G543D, G543V, L547H, L547P, L547F, L547R, C537Y, M548I, M548L, M548K, |

M548R, M548T, G542V,G550R, C554Y, R556K, R556S, R556G, C559R, C559Y, C562Y, V563M, P580L, W581R, W581G, W581C, W581S, C583S, C583Y, C586G, C586R, C586Y, K589N, L595P, R596W, R598Q, R604Q, R604W, P633H, P633L, I634F, I634T, R635G, R635L, R635P, R635W, R635Q, V636A, V636G, V636M, V636L, L637R, L637P, L637Q, S638F, S638P, S638Y, L639R, L639V, L639F, A644T, T645A, G646V, G646E, L647H, L648P, V649G, V649L, V649M, L650V, L650Q, L653W, L653F, I655N, I655T, Q656K, V657A, V657M, V657G, D658V, D658Y, R659C, R659G, R659H, Y660C, Y660N, Y660F, Y660H, Y660D, A662D, S663L, S663W, E664K, V665G, V665L, S669F, S669P, M674V, V675A, V675M, R676L, R676W, R676Q, I681N, I681S, I681M, M682R, Y683D, V684F, G685R, G685E, G685A, D686Y, D686G, D686H, D686V, V687L, D686A, V687F, R688C, R688G, R688H, V690G, V690F, V690D, T691I, I695N, H694Y, H694P, I695F I695T, Q696P,W698C, W698R, W698S G699R, G699S, G699D, G699V, P700L, P700S, P700R, P700Q, P700T, P700A, F701V, D702A, D702G. D702E, D702V, D702N, D702Y, L703P, L703R, L703V, V704A, V704M, V704G, I705F, I705T, I705S, I705N, G706E, G706W, G706R, G706V, G707C, G707D, G707S, G707R, G707V, C710S, C710Y, D712A, L713F, S714C, V716D, V716F, V716I, N717S, N717I, P718L, R720C, R720H, R720G, R720S, K721R, K721T, K721N, Y724C, E725K, G726V, G728D, R729Q, R729W, R729G, R729L, F731C, F731L, F731Y, F731I, F731V, F732del, F732C, F732I, F732S, F732L, F732V, E733G, E733A, E733V, F734L, F734C, F734V, Y735C, Y735N, Y735S, Y735F, Y735H, R736G, R736H, R736C, R736L, R736P, R736S, L737H, L737P, L737V, L737F, L737R, L737P, L738P, L738Q, H739P, A741G, A741V, R742L, R742G, R742P, P743H, P743R, P743L, P743S, R749C, R749L, R749H, R749G, P750R, F751L, F751C, F751I, F751V, F752del, F752C, F752L, F752I, F752V, W753G, W753C, W753L, W753R, W753S, L754P, L754R, L754H, F755S, F755I, F755L, M761I, M761V, G762C, V763G, V763I, K766E, D768E, D768H, D768V, D768Y, I769N, I769S, I769T, I769V, S770L, S770W, S770P, R771G, R771L, R771P, R771Q, F772C, F772I, F772V, L773H, L773I, L773R, L773V, E774A, E774K, E774D, E774G, E774V, S775F, S775P, P777A, P777H, P777L, P777R, P777T, P777S, V778M, I780N, I780S, I780T, D781G, V785M, A787G, A787S, H789Q, R790W, A791V, R792C, R792H, R792S, F794L, F794V, W795S, W795G, W795C, W795L, W795R, G796A, G796C, G796D, G796V, N797D, N797Y, N797H, L798P, L798H, N797K, N797S, P799L, P799A, P799T, P799S, P799R, P799H, G800S, M801I, M801R, M801T, M801V, R803S, R803G, R803K, R803T, R803W, R803M, P804L, P804S, L815Q, H821D, H821P, H821R, K826N, K826T, K826R, S828N, K829R, T835M, N838D, S839P, K841N, K841T, K841Q, Q842E, Q842R, G843S, P849L, M853R, M852L, M852V, D857N, W860R, E863D, E863G, E863K, E863V, F868S, F868L, G869S, G869V, F870V, H873R, Y874C, R879D, M880I,

| | |
|---|---|
| | M880L, M880V, S881R, S881I, R882H, R882P, R882L, R882C, R882G, R882S, A884P, A884V, R885K, Q886E, Q886R, L889P, L889R, G890D, G890R, G890S, G890V, W893S, V895M, P896L, P898S, V897G, V897D, I898T, R899P, R899L, R899G, R899H, R899C, R899S, L901P, L901R, L901H, L901V, A903P, A903T, P904L, P904Q, P904A, P904R, P904S, L905R, L905P, L905Q, L905V, K906E, E907G, Y908C, Y908D, Y908N, F909C, A910P, A910V, C911R, C911Y |
| ***EED*** | Frameshift/nonsense, L240Q, I363M |
| ***EP300*** | Frameshift/nonsense, VF1148_1149del, D1399N, D1399Y, P1452L, Y1467N, Y1467H, Y1467C, R1627W, A1629V |
| ***ETNK1*** | N244S, N244T, N244K |
| ***ETV6*** | Frameshift/nonsense |
| ***EZH2*** | Frameshift/nonsense, Q62R, N102S, F145S, F145C, F145Y, F145L, G159R, E164D, R202Q, K238E, E244K, R283Q, H292R, P488S, R497Q, R561H, T568I, K629E, Y641N, Y641H, Y641S, Y641C, Y641F, D659Y, D659G, V674M, A677G, A677V, R679C, R679H, R685C, R685H, A687V, N688I, N688K, H689Y, S690P, I708V, I708T, I708M, E720K, E740K |
| ***FLT3*** | V579A, V592I, F594L, FY590-591GD, D835Y, D835H, D835E, del835 |
| ***GATA1*** | Frameshift/nonsense |
| ***GATA2*** | Frameshift/nonsense, R293Q, N317H, A318T, A318V, A318G, G320D, L321P, L321F, L321V, Q328P, R330Q, R361L, L359V, A372T, R384G, R384K |
| ***GATA3*** | Frameshift/nonsense/splice-site ZNF domain, R276W, R276Q, N286T, L348V |
| ***GNA13*** | I34T, G57S, S62F, M68K, Q134R, Y145F, L152F, E167D, Q169H, R264H, E273K, V322G, V362G, L371F |
| ***GNAS*** | R201(844) G, R201(844) S, R201(844) C, R201(844) H, R201(844)L, Q227(870)K, Q227(870)R, Q227(870)L, Q227(870)H, R374(1017)C |
| ***GNB1*** | K57N, K57M, K57E, K57T, I80T, I80N |
| ***IDH1*** | R132C, R132G, R132H, R132L, R132P, R132V |
| ***IDH2*** | R140W, R140Q, R140L, R140G, R172W, R172G, R172K, R172T, R172M, R172N, R172S |
| ***IKZF1*** | Frameshift/nonsense |
| ***IKZF2*** | Frameshift/nonsense |
| ***IKZF3*** | Frameshift/nonsense |
| ***JAK1*** | T478A, T478S, V623A, A634D, L653F, R724H, R724Q, R724P, T782M, L783F |
| ***JAK2*** | N533D, N533Y, N533S, H538R, K539E, K539L, I540T, I540V, V617F, R683S, R683G, del/ins537-539L, del/ins538-539L, del/ins540-543MK, del/ins540-544MK, del/ins541-543K, del542-543, del543-544, ins11546-547 |

| | |
|---|---|
| ***JAK3*** | M511T, M511I, A572V, A572T, A573V, R657Q, V715I, V715A |
| ***KDM6A*** | Frameshift/nonsense/splice-site, del419 |
| ***KIT*** | ins503, V559A, V559D, V559G, V559I, V560D, V560A, V560G, V560E, del560, E561K, del579, P627L, P627T, R634W, K642E, V654A, V654E, H697D, E761D, K807R, D816H, D816Y, D816F, D816I, D816V, D816H, del551-559 |
| ***KRAS*** | G12D, G12A, G12E, G12V, G13D, G13C, G13Y, G13F, G13R, G13A, G13V, G13E, V14I, L19F, T58I, G60D, G60A, G60V, Q61K, Q61E, Q61P, Q61R, Q61L, Q61H, K117E, K117N, A146T, A146P, A146V |
| ***LUC7L2*** | Frameshift/nonsense/splice-site |
| ***MLL*** | Frameshift/nonsense |
| ***MLL2*** | Frameshift/nonsense |
| ***MPL*** | S505G, S505N, S505C, L510P, del513, W515A, W515R, W515K, W515S, W515L, A519T, A519V, Y591D, W515-518KT |
| ***NF1*** | Frameshift/nonsense |
| ***NPM1*** | Frameshift p. W288fs (insertion at c.859_860, 860_861, 862_863, 863_864) |
| ***NRAS*** | G12S, G12R, G12C, G12N, G12P, G12Y, G12D, G12A, G12V, G12E, G13S, G13R, G13C, G13N, G13P, G13Y, G13D, G13A, G13V, G13E, G60E, G60R, Q61R, Q61L, Q61K, Q61P, Q61H, Q61Q |
| ***PDS5B*** | Frameshift/nonsense/splice-site, R1292Q |
| ***PDSS2*** | Frameshift/nonsense |
| ***PHF6*** | Frameshift/nonsense/splice-site, A40D, M125I, S246Y, F263L, R274Q, C297Y, H302Y, H329L |
| ***PHIP*** | Frameshift/nonsense/splice-site |
| ***PPM1D*** | Frameshift/nonsense, exon 5 or 6 |
| ***PRPF40B*** | Frameshift/nonsense/splice-site, P15H, M58I, P405L, P562S, |
| ***PRPF8*** | M1307I, C1594W, D1598Y, D1598N, D1598V (ADD MORE VARS) |
| ***PTEN*** | Frameshift/nonsense, D24G, R47G, F56V, L57W, H61R, K66N, Y68H, C71Y, F81C, Y88C, D92G, D92V, D92E, H93Y, H93D, H93Q, N94I, P95L, I101T, C105F, C105S, D107Y, L112V, H123Y, C124R, C124S, K125E, A126D, K128N, R130G, R130Q, R130L, G132D, I135V, I135K, C136R, C136F, K144Q, A151T, D153Y, D153N, Y155H, Y155C, R159K, R159S, R161K, R161I, G165R, G165E, S170N, S170I, R173C, Y174D, Y177C, H196Y, R234W, G251C, D252Y, F271S, D326G |
| ***PTPN11*** | G60V, G60R, G60A, D61Y, D61V, D61G, Y63C, E69K, E69G, E69D, E69Q, F71L, F71K, A72T, A72V, A72D, T73I, E76K, E76Q, E76M, E76A, E76G, E139G, E139D, N308D, N308T, N339S, P491L, S502P, S502A, S502L, G503V, G503G, G503A, G503E, Q506P, T507A, T507K |

| | |
|---|---|
| ***RAD21*** | Frameshift/nonsense/splice-site, R65Q, H208R, Q474R |
| ***RUNX1*** | Frameshift/nonsense/splice-site, S73F, H78Q, H78L, R80C, R80P, R80H, L85Q, P86L, P86H, S114L, D133Y, L134P, R135G, R135K, R135S, R139Q, R142S, A165V, R174Q, R177L, R177Q, A224T, D171G, D171V, D171N, R205W, R223C |
| ***SETBP1*** | D868N, D868T, S869N, G870S, I871T, D880N, D880Q |
| ***SETD2*** | Frameshift/nonsense, V1190M |
| ***SETDB1*** | Frameshift/nonsense, K715E |
| ***SF1*** | Frameshift/nonsense/splice-site, T454M, Y476C, A508G |
| ***SF3A1*** | Frameshift/nonsense/splice-site, A57S, M117I, K166T, Y271C |
| ***SF3B1*** | G347V, R387W, R387Q, E592K, E622D, Y623C, R625L, R625C, R625G, N626D, R630G, H662Q, H662D, T663I, K666N, K666Q, K666T, K666E, K666R, K700E, V701F, A708T, G740R, G740E, G742D, A744P, A745P, K748E, R775P, D781G, E783K, R831Q, L833F, E862K, R957Q |
| ***SFRS2*** | Y44H, P95H, P95L, P95T, P95R, P95A, P107H, P95fs |
| ***SMC1A*** | K190T, R586W, M689V, R807H, R1090H, R1090C |
| ***SMC3*** | Frameshift/nonsense, R155I, Q367E, D392V, K571R, R661P, G662C |
| ***STAG1*** | Frameshift/nonsense/splice-site, H1085Y |
| ***STAG2*** | Frameshift/nonsense/splice-site |
| ***SUZ12*** | Frameshift/nonsense |
| ***TET2*** | Frameshift/nonsense/splice-site, missense mutations in catalytic domains (p.1104-1481 and 1843-2002), D1121Y, D1129Y, C1133R, C1133W, C1135Y, C1135F, C1135W, G1137D, G1137V, E1137K, E1137D, E1141K, E1144K, Y1148C, L1151R, G1152R, A1153T, A1153V, G1154S, C1156Y, V1157M, I1160F, I1160S, R1161G, R1161S, M1164I, E1165K, E1165D, R1167G, R1167K, R1167S, R1167M, L1172R, A1174T, I1175T, V1180D, M1185I, E1186A, G1187S, K1188R, G1192V, C1193Y, C1193W, P1194L, P1194R, I1195V, K1197E, W1198C, W1198R, V1201I, E1207D, L1209P, L1210P, C1211Y, L1212S, V1213M, V1213E, R1214W, R1214Q, R1216Q, H1219D, H1219R, H1219Y, C1221Y, C1221R, C1221S, C1221W, C1221F, L1229R, G1235E, R1235W, L1238V, A1241S, K1243R, K1243N, L1244P, Y1245C, Y1245N, L1248P, L1248R, L1252P, L1252V, G1256C, R1261C, R1261S, R1261H, R1261P, R1262W, C1263F, C1263Y, N1266D, N1266K, N1266H, N1266Y, N1266S, C1271W, C1271S, C1273S, C1273W, C1273R, Q1274P, G1275R, G1275V, G1282R, G1282D, R1283P, S1284F, F1287V, G1288D, G1288V, C1289F, C1289W, S1290L, W1291C, S1292R, M1293I, Y1294C, G1297E, G1297R, C1298Y, C1298S, K1299M, K1299Q, K1299N, F1300V, F1300L, F1300I, S1303G, S1303R, K1310Q, L1311Q, E1318E, L1322Q, L1322P, L1322R, L1326W, L1329P, L1329Q, L1332P, |

M1333K, L1340R, L1340P, Y1345D, Y1345C, Q1348K, Q1348R, I1349N, E1352K, A1355V, C1358S, C1358W, R1359C, R1359L, R1359P, R1359H, R1359G, R1359S, L1360R, G1361C, G1361S, G1361D, R1366H, R1366L, R1366C, P1367L, P1367S, P1367R, F1368L, G1370V, G1370R, V1371D, A1373P, A1376V, D1376G, F1377V, F1377I, C1378R, C1378Y, C1378F, H1380Y, H1380L, H1380R, H1380Q, R1380C, H1380D, H1382R, H1382P, R1383G, H1386D, R1387H, N1387S, T1393A, C1395Y, T1397I, L1398R, F1398C, F1398L, L1398H, L1398P, H1401Y, E1401A, N1403S, Q1414R, Q1414H, Q1414K, V1416I, V1417F, P1419R, D1427V, Q1435K, V1438F, G1861E, G1861R, G1861V, A1863V, H1868Y, H1868P, H1868L, G1869W, S1870L, L1872P, I1873T, I1873S, A1876T, A1876V, R1878P, R1878H, E1879Q, E1879G, E1879D, H1881N, H1881L, T1884A, T1884I, P1889L, N1890S, P1894T, P1894R, P1894L, P1894H, R1896G, I1897N, S1898F, V1900F, E1900K, V1900D, Y1902H, Y1902C, Q1903R, A1903V, H1904R, H1904Q, K1905E, T1905A, H1912R, H1912D, H1912Y, G1913D, A1919D, F1922S, Y1923H, H1925Q, K1934N, R1966C, D1981A, S1982Y

***TP53*** Frameshift/nonsense/splice-site, S46F, G105C, G105R, G105D, G108S, G108C, R110L, R110C, T118A, T118R, T118I, S127F, S127Y, L130V, L130F, K132Q, K132E, K132W, K132R, K132M, K132N, F134V. F134L, F134S, C135W, C135S, C135F, C135G, C135Y, Q136K, Q136E, Q136P, Q136R, Q136L, Q136H, A138P, A138V, A138A, A138T, T140I, C141R, C141G, C141A, C141Y, C141S, C141F, C141W, V143M, V143A, V143E, L145Q, W146C, W146L, L145R, V147G, P151T, P151A, P151S, P151H, P151R, P152S, P152R, P152L, T155P, T155A, V157F, R158H, R158L, A159V, A159P, A159S, A159D, A161T, A161D, Y163N, Y163H, Y163D, Y163S, Y163C, K164E, K164M, K164N, K164P, H168Y, H168P, H168R, H168L, H168Q, M169I, M169T, M169V, E171K, E171Q, E171G, E171A, E171V, E171D, V172D, V173M, V173L, V173G, R174W, R175G, R175C, R175H, C176R, C176G, C176Y, C176F, C176S, P177R, P177L, H178D, H178P, H178Q, H179Y, H179R, H179D, H179Q, R181C, R181Y, R181H, D186G, G187S, P190L, P190T, H193N, H193P, H193L, H193R, L194F, L194R, I195F, I195N, I195T, R196P, V197L, G199V, Y205D, Y205N, Y205C, V203M, Y205H, D208V, R213Q, R213P, F212I, R213L, R213Q, H214D, H214P, H214R, S215G, S215I, S215R, V216M, V217G, Y220N, Y220H, Y220S, Y220C, E224D, I232F, I232N, I232T, I232S, Y234N, Y234H, Y234S, Y234C, Y236N, Y236H, Y236C, M237V, M237K, M237I, C238R, C238G, C238Y, C238W, N239T, N239S, S241Y, S241C, S241F, C242G, C242Y, C242S, C242F, G244S, G244C, G244D, G245S, G245R, G245C, G245D, G245A, G245V, G245S, M246V, M246K, M246R, M246I, N247I, R248W, R248G, R248Q, R249G, R249W, R249T, R249M, P250L, I251N, L252P, I254S, I255F, I255N, I255S, L257Q, L257P, E258K, E258Q, D259Y, S261T, G262D, G262V, L265P, G266R, G266E, G266V, R267W, R267Q, R267P,

| | |
|---|---|
| | E271K, V272M, V272L, R273S, R273G, R273C, R273H, R273P, R273L, V274F, V274D, V274A, V274G, V274L, C275Y, C275S, C275F, A276P, C277F, C277Y, P278T, P278A, P278S, P278H, P278R, P278L, G279E, R280G, R280K, R280T, R280I, R280S, D281N, D281H, D281Y, D281G, D281E, D281V, R282G, R282W, R282Q, R282P, E285K, E285V, E286G, E286V, E286K, K320N, L330R, G334V, R337C, R337L, A347T, L348F, T377P |
| ***U2AF1*** | D14G, S34F, S34Y, R35L, R156H, R156Q, Q157R, Q157P |
| ***U2AF2*** | R18W, Q143L, M144I, L187V, Q190L |
| ***WT1*** | Frameshift/nonsense/splice-site |
| ***ZRSR2*** | Frameshift/nonsense, E133G, C181F, D185G, C187Y, H191Y, I202N, F239V, F239Y, N261Y, C280R, C302R, C326R, H330R |

**Supplemental Table 8. Billing codes used to define outcomes.**

| Outcome | ICD Codes | UK Biobank data-fields |
|---|---|---|
| **Primary outcome (any CVD event)** | Any from “MI”, “CAD”, “Stroke”, or “Death” below | Any from “MI”, “CAD”, “Stroke”, or “Death” below |
| **MI** | | 42001 (values: 1,2) |
| **CAD (Revascularization)** | K40.1, K40.2, K40.3, K40.4, K41.1, K41.2, K41.3, K41.4, K45.1, K45.2, K45.3, K45.4, K45.5, K49.1, K49.2, K49.8, K49.9, K50.2, K75.1, K75.2, K75.3, K75.4, K75.8, K75.9 | 42001 (values: 1,2) |
| **Stroke** | | 42007 (values: 1,2) |
| **Death** | | 40020 (any value) |
| **Type 2 Diabetes** | E11, E11.0, E11.1, E11.2, E11.3, E11.4, E11.5, E11.6, E11.7, E11.8, E11.9 | 20002 (values: 1223) |

CVD: cardiovascular disease, MI: myocardial infarction

**IL6R** | **Il1B** | **NLRP3**

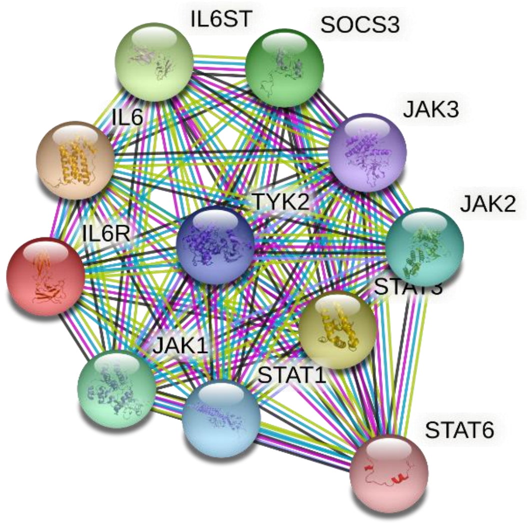


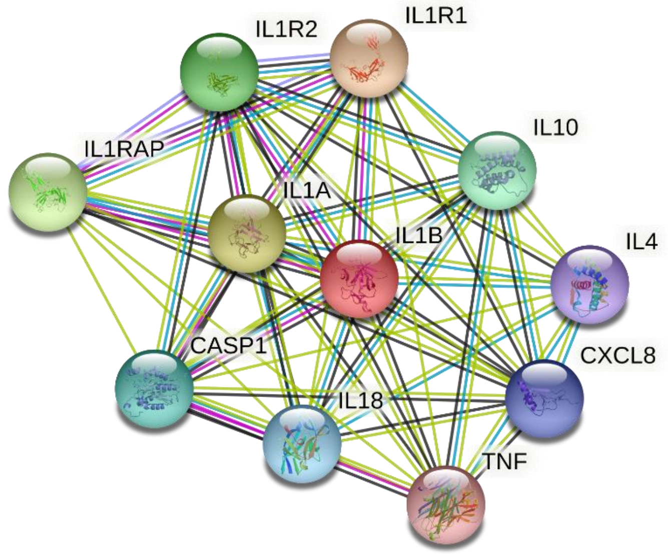


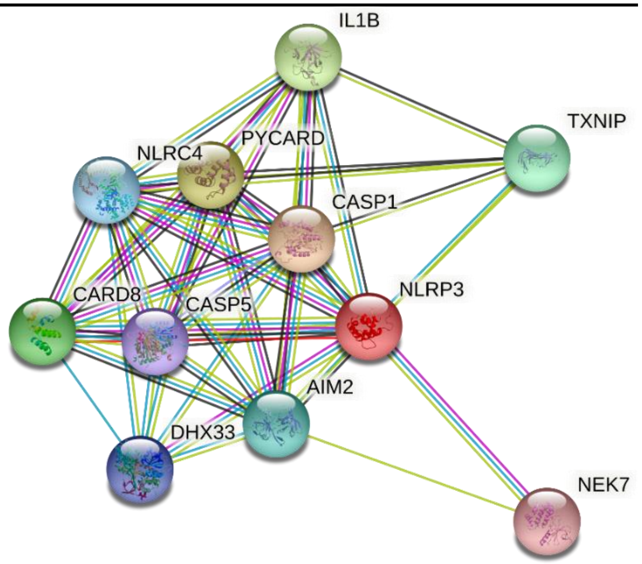


**IFNG** | **IL18**

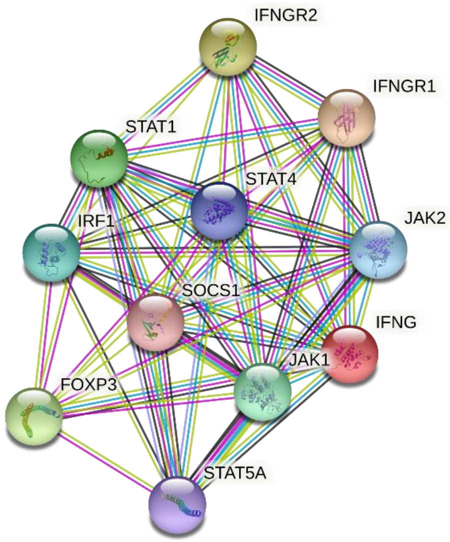


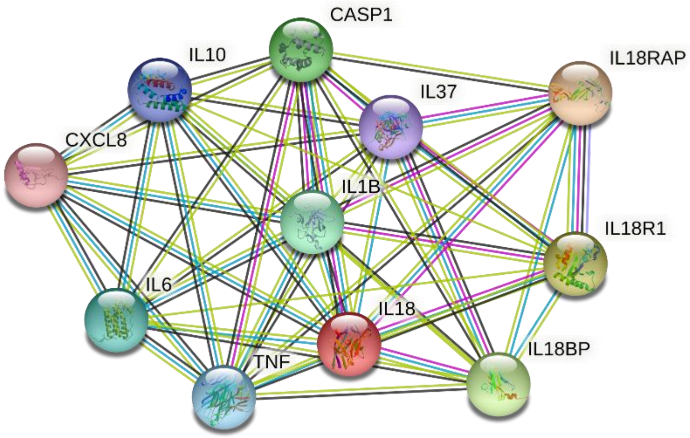

**Supplemental Figure 1. Protein-protein interaction network of IL-6R, IL-1B, NLRP3, IFNG, and IL-18.** Network structures were obtained from the STRING database (v11; https://string-db.org/). The top ten proteins that interact with IL-6R, IL-1B, NLRP3, IFNG, and IL-18 were presented and selected.

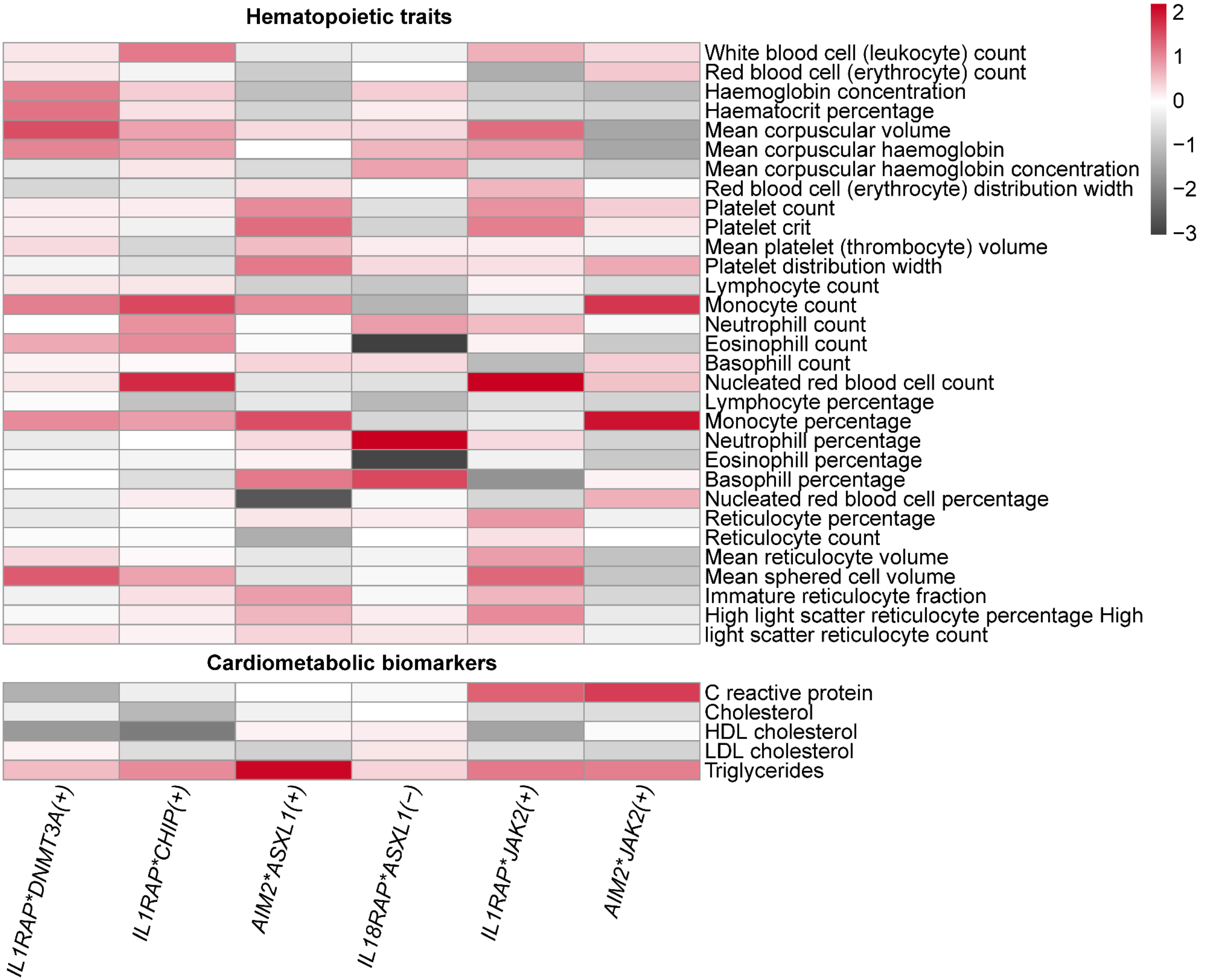
Hematopoietic traits
White blood cell (leukocyte) count
Red blood cell (erythrocyte) count
Haemoglobin concentration
Haematocrit percentage
Mean corpuscular volume
Mean corpuscular haemoglobin
Mean corpuscular haemoglobin concentration
Red blood cell (erythrocyte) distribution width
Platelet count
Platelet crit
Mean platelet (thrombocyte) volume
Platelet distribution width
Lymphocyte count
Monocyte count
Neutrophill count
Eosinophill count
Basophill count
Nucleated red blood cell count
Lymphocyte percentage
Monocyte percentage
Neutrophill percentage
Eosinophill percentage
Basophill percentage
Nucleated red blood cell percentage
Reticulocyte percentage
Reticulocyte count
Mean reticulocyte volume
Mean sphered cell volume
Immature reticulocyte fraction
High light scatter reticulocyte percentage High light scatter reticulocyte count
Cardiometabolic biomarkers
C reactive protein
Cholesterol
HDL cholesterol
LDL cholesterol
Triglycerides
IL1RAP*DNMT3A(+)
IL1RAP*CHIP(+)
AIM2*ASXL1(+)
IL18RAP*ASXL1(−)
IL1RAP*JAK2(+)
AIM2*JAK2(+)
2
1
0
−1
−2
−3

**Supplemental Figure 2. Heatmap for the Z-scores of associations between predicted expression scores of inflammatory genes and hematopoietic traits and cardiometabolic biomarkers in the presence of CHIP mutations.** For CHIP mutation-predicted expression score pairs that showed significant interaction for CVD event incidence, we examined the association between the score of inflammatory gene and hematopoietic traits and cardiometabolic biomarkers among participants with the corresponding CHIP mutations. Predicted expression scores of inflammatory genes were calculated by applying either P+T or PRS-CS methods to the summary statistics of the eQTL for those genes from the eQTLGen Consortium (https://www.eqtlgen.org/)**.** Black color indicates a negative Z-score, and red indicates a positive Z-score. No association passed FDR<0.05 or 0.1 level. The darker the color, the stronger the effects. CHIP, clonal hematopoiesis of indeterminate potential. eQTL: expression quantitative trait loci; PRS-CS: polygenic risk score-continuous shrinkage; P+T: pruning and thresholding.
